# A controlled human infection model for symptomatic pertussis in North America using the pertactin-producing clinical isolate D420

**DOI:** 10.64898/2026.06.09.26355227

**Authors:** May S ElSherif, Kara L Redden, Joanne M Langley, Lingyun Ye, Wade Blanchard, Bruce Smith, Jun Wang, Bahaa Abu-Raya, Jillian H Filliter, Kathryn M Edwards, C. Buddy Creech, Shelly McNeil, Todd F Hatchette, Jason J LeBlanc, Susan Hariri, Lucia Pawloski, Panagiotis Maniatis, LeAnne M Fox, Carrie A Whittle, Scott A Halperin

## Abstract

**Background:** Despite widespread vaccination, pertussis remains a poorly controlled disease globally and results in substantial annual morbidity and mortality, particularly in young children. Controlled human infection models (CHIMs) using the causative agent *Bordetella pertussis* are promising systems to enable the study of pertussis disease pathogenesis and immunology and to rapidly assess vaccines and therapeutics. While a pertussis CHIM that produces asymptomatic infection has been established in Europe, the development of a CHIM that leads to symptomatic illness would be advantageous for evaluating vaccine efficacy against both infection and disease.

**Methods:** Healthy participants 18–40 years of age were inoculated intranasally with one of eight doses (ranging from 10^4^ to 10^8^ colony forming units (CFU)) of the pertactin-producing *B. pertussis* isolate D420 at the challenge facility within the Canadian Center for Vaccinology (Nova Scotia, Canada). The study occurred in two stages. In stage one, the *B. pertussis* dose was escalated in cohort groups of five to six participants until reaching an endpoint where 70-90% of participants exhibited mild (non-severe, Grade 1 or 2) symptomatic infection, defined as the Human Infectious Dose 70-90 (HID70-90). In stage two, additional challenges were conducted for doses below, at, and above the identified HID70-90 to characterize the emerging pertussis model. For all challenge doses, participants were closely monitored during an inpatient stay of up to 24 days and post-discharge for laboratory-confirmed infection, pertussis symptoms, safety, and IgG antibody responses to four *B. pertussis* antigens including pertussis toxin, filamentous hemagglutinin, fimbriae, and pertactin. All participants received a five-day course of azithromycin, where timing of initiation depended on *B. pertussis* testing and symptoms. The study was conducted between July 4, 2022 and March 19, 2025.

**Findings:** Seventy-five participants were inoculated with one of the eight *B. pertussis* D420 challenge doses and completed the inpatient stay. From the stage-one dose escalation, we found that 10^7^ CFU of *B. pertussis* D420 was the lowest dose that achieved the HID70-90, where 9 of 12 participants (75·0%) exhibited mild symptomatic infection. Following stage-two challenges, 16 of 22 total participants at 10^7^ CFU (72·7%) developed mild symptomatic infection, thus verifying the HID70-90. The symptomatic infection rate below the HID70-90 at 5×10^6^ CFU of D420 was 20·0% and above the HID70-90 at 5×10^7^ and 10^8^ CFU were 58·3% and 55·6%, respectively. Symptoms with elevated frequency for symptomatic infection (relative to background symptoms in non-infected) included nasal congestion, runny nose, fatigue, malaise, and cough. At the HID70-90, 50% of symptomatic infections included cough. Serological analyses of the four highest (stage-two) challenge doses (5×10^6^, 10^7^, 5×10^7^, 10^8^ CFU) revealed that antibody titres increased over time post-challenge. Seroconversion for at least one of the four studied antibodies was nearly twice as common for symptomatic (70·0%) than asymptomatic (35·7%) infection and was absent (0%) for non-infected. All infections were cleared following azithromycin treatment (100%) and there were no study-related serious adverse events.

**Interpretation:** A safe and reproducible symptomatic pertussis CHIM was achieved, providing a model for research on pertussis disease pathogenesis and immunology and for assessing vaccines and therapeutics. (Clinicaltrials.gov, NCT05136599).

**Funding:** United States (US) Centers for Disease Control and Prevention, US National Institutes of Health

**Research in Context:** *Evidence before this study:* *Bordetella pertussis*, the causative agent of pertussis (whooping cough), remains a global threat despite nearly universal vaccination programs. The burden of pertussis disease is highest for infants but can also lead to complications among some adults, particularly the elderly and immunocompromised. Suboptimal vaccine efficacy and waning immunity post-vaccination have led to resurgences of pertussis in many countries and numerous outbreaks. New strategies are needed to investigate the pathophysiology and immunology of *B. pertussis* infection and to develop more efficacious vaccines and therapeutics. Controlled human infection models (CHIMs), the intentional exposure of human volunteers to a pathogen under closely monitored conditions, are promising platforms to address knowledge gaps on the disease and to assess the efficacy of novel vaccines. Recently, through a European collaborative effort, an asymptomatic pertussis CHIM was established using the *B. pertussis* isolate B1917 that showed intranasal bacterial challenge doses of 10^4^ to 10⁵ colony-forming units (CFU) produced infection and antibody responses, was safe, and could be eliminated with antibiotics. The development of a symptomatic pertussis CHIM would permit evaluation of vaccine efficacy against both infection and symptomatic disease, and better reflect real-world circumstances.

*Added value of this study:* This study describes the establishment of a pertussis CHIM that induces mild symptomatic infection using the pertactin-producing isolate D420 at the challenge unit facilities of the Canadian Center for Vaccinology (Nova Scotia, Canada). Infection progression for participants in the study was closely monitored by testing for the pathogen in nasal washes (cultures and polymerase chain reaction (PCRs)), symptom assessments, and assays of pertussis antibodies. Using a dose-escalation design for nasal challenge with D420, we found that a *B. pertussis* dose of 10^7^ CFU led to symptomatic disease in over 70% of participants. Symptoms associated with symptomatic infection included nasal congestion, runny nose, fatigue, malaise, and cough. Additional challenges below, at, and above the HID70-90 (5×10^6^, 10^7^, 5×10^7^, 10^8^ CFU) provided insights into infection rates, symptom development, antibody responses, and safety of the CHIM. Overall, all symptoms were low-grade (Grade 1 or 2), infections were cleared with azithromycin, and there were no study-related serious adverse events. We demonstrate that different rates of asymptomatic and symptomatic infection were safely achieved by titrating the challenge dose of *B. pertussis*.

*Implications of all the available evidence:* The pertussis CHIM described here provides a platform to evaluate efficacy and safety of novel vaccine candidates and therapeutics against pertussis infection and disease. The CHIM also provides a framework to advance fundamental research in pertussis disease onset and progression, including correlates of infection, symptomology, and immune protection.

## Introduction

Pertussis (whooping cough) remains poorly controlled globally despite widespread vaccination.^1,2^ The disease burden is highest among infants (∼400,000 annual deaths globally),^2^ and can give rise to complications among some adults including the elderly and those with chronic illness.^3^ Unlike some infectious diseases, long-term immunity to the causative bacterial agent, *Bordetella pertussis*, is not sustained after natural infection or by vaccination with whole cell pertussis (wP) or acellular pertussis (aP) vaccines.^1,2,4,5^ Innovative research strategies are needed to better understand pertussis infection, disease, pathogenesis, and immune responses, and to evaluate novel vaccines and therapeutics.^4,6,7^

A controlled human infection model (CHIM) provides a versatile platform to study infectious disease.^7,8^ In a CHIM, human participants are intentionally exposed to a pathogen in a controlled, safe, and medically-supervised environment, allowing close-up research on disease progression, pathogenesis, and immune responses, and rapid assessment of the efficacy and safety of vaccines and therapeutics.^8,9^ A pertussis CHIM permits investigation of *B. pertussis* infection in its only natural (human) host,^2,10^ and overcomes limitations of research in animal models (e.g., mouse, baboon), which do not replicate the spectrum of symptoms or clinical phenotypes of humans.^11–13^ Further, infection progression may be studied at regular intervals, at specific pathogen doses, and with close monitoring of symptoms including cough.^2,6,14^ Unlike population-level pertussis research, which has focused primarily on the period after the symptoms and the distinctive paroxysmal cough have emerged,^6,15,16^ a pertussis CHIM is suitable for studying early phases of the infection and disease. Overall, the development of pertussis CHIMs has the potential to expand our research capacity on pertussis and to reduce our reliance on large-scale, costly, multi-year trials for evaluating vaccine efficacy and safety.^13^

Recently, an asymptomatic pertussis CHIM was established in Europe using the pertactin-producing isolate B1917 wherein intranasal inoculation at doses of 10^4^ to 10⁵ colony-forming units (CFU) resulted in infection and serological responses without significant safety events.^7,8^ Notwithstanding this advance, there is an eminent need for a pertussis CHIM that induces symptomatic infection which more closely reflects clinical disease in real-world conditions.^2,3^ Here, we report the establishment of a symptomatic pertussis CHIM in healthy adults using the *B. pertussis* pertactin-producing clinical isolate D420, a wild-type strain originating from North America.^17–19^

## Methods

### Study objectives

The primary objective of this study was to develop and characterize a symptomatic pertussis CHIM. We aimed to identify a safe intranasal Human Infectious Dose (HID) of *B. pertussis* D420 that reproducibly induced mild symptomatic infection at an attack rate of 70-90% (HID70-90). The study was an open-label phase 1 dose escalation trial (without placebo) conducted in the challenge unit facility at the Canadian Center for Vaccinology (CCfV) (Halifax, Nova Scotia, Canada) between July 4^th^ 2022 and March 19, 2025. The primary endpoints included laboratory-confirmed infection (using culture and/or polymerase chain reaction (PCR) testing of nasal wash) post-challenge with reporting of mild pertussis-related symptoms. Secondary endpoints included post-challenge production of the antibodies immunoglobin G (IgG) anti-pertussis toxin (anti-PT), filamentous hemagglutinin (anti-FHA), fimbriae (anti-FIM), and pertactin (anti-PRN) up to 56 days post-challenge.

### Participants

Healthy adults aged 18–40 years were eligible to participate if they had no history of laboratory-confirmed pertussis, had not received a pertussis vaccine within the previous five years and not more than seven cumulative pertussis vaccine doses since infancy. Additional inclusion criteria included PT IgG antibody levels ≤20 enzyme-linked immuno-absorbent assay (ELISA) units per mL (IU/mL) and nasal samples negative for *B. pertussis* by culture and PCR at screening. Inclusion and exclusion criteria are provided in Appendix A.

### Study Design

The study occurred in two stages. The first stage employed a dose-escalation design in sequential cohorts of five to six participants to determine the lowest intranasal dose of *B. pertussis* D420 that safely produced mild symptomatic infection in 70–90% of participants (HID70–90). Based on safety data reported in prior research,^8^ the initial cohort group in the study was challenged with a single 10^4^ CFU dose, with subsequent cohorts receiving 2-, 5- or 10-fold (2x, 5x or 10x) dose escalations until the target symptomatic infection rate (HID70-90) was achieved. The first dose that led to the putative HID70-90 was then repeated in another cohort group to assess reproducibility.

The second stage involved additional challenges to evaluate the emerging model across four dose levels: 1) 0.5x the HID70-90; 2) the HID70-90; 3) 5x above the HID70-90; and 4) 10x the HID70-90 (figure 1). The objective was to further characterize symptomatic infection rates in the emerging CHIM including nasal *B. pertussis* levels (load), symptom profiles, antibody responses, and safety (figure 1).

**Figure 1.**
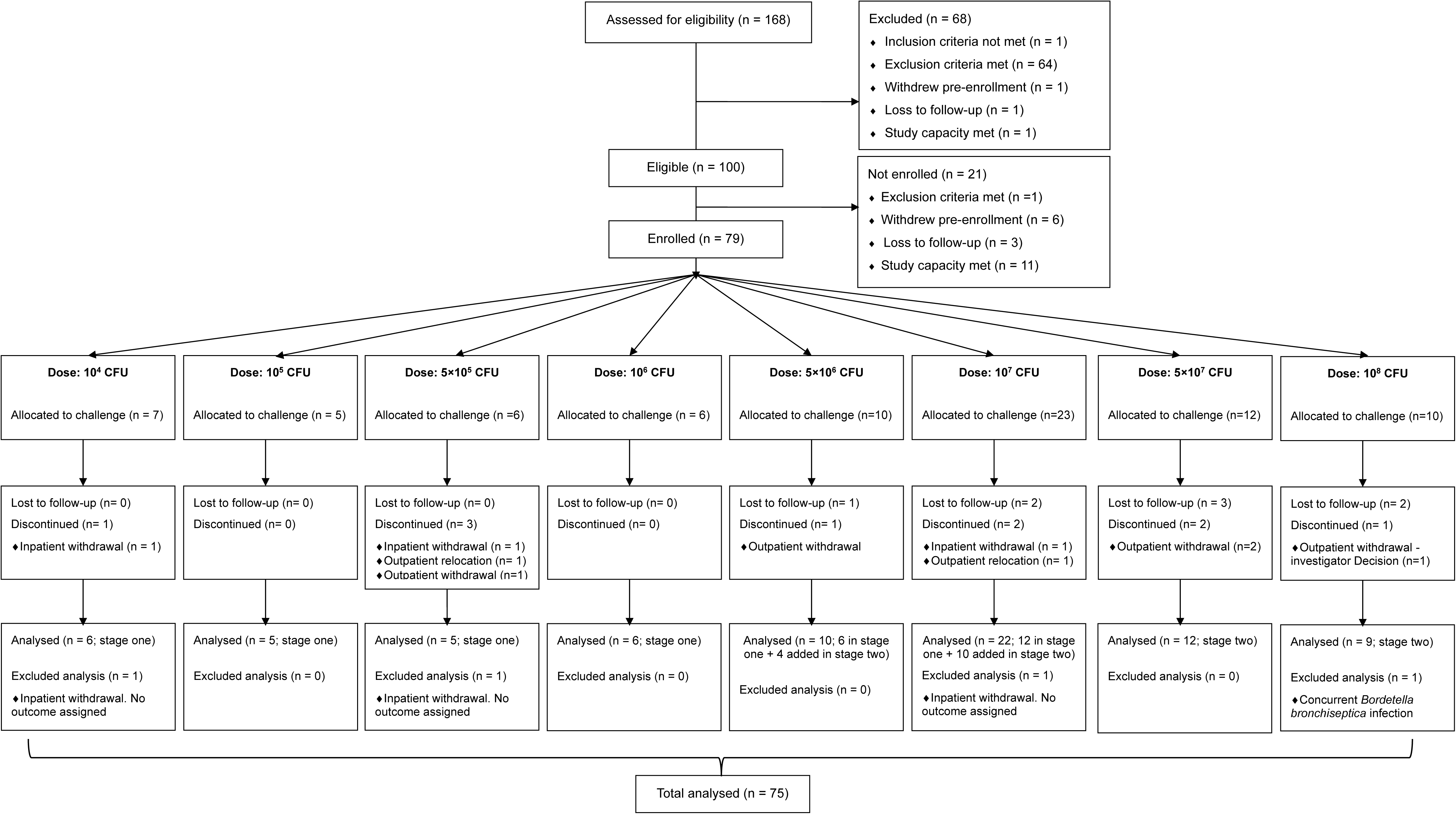
CONSORT flow diagram showing the total number (n) of participants challenged and analysed per challenge dose.

### Data Safety Monitoring Board

An independent Data Safety Monitoring Board (DSMB) evaluated the safety data for each cohort group once during the inpatient stay to decide whether the challenge dose could be escalated in the next cohort, and once again during the outpatient period. For the two highest doses under study (figure 1), a sentinel challenge approach was used; three participants were challenged and evaluated by the DSMB for safety approval before the remaining participants were challenged.

### Ethics

This study was conducted in accordance with the Declaration of Helsinki and the International Conference on Harmonisation guidelines for Good Clinical Practice (ICH GCP) and approved by the IWK Health’s Research Ethics Board (Institutional Review Board #IRB00003719, FWA #FWA00005630, IORG #IORG000310). All participants gave written informed consent prior to enrollment and study procedures.

### Challenge material and administration

The *B. pertussis* D420 challenge material was manufactured at CCfV using Good Manufacturing-like Practices (GMP-like). The live-GMP-like liquid culture was aliquoted and stored at −70°C (±10°C). A single aliquot was used per participant. On the day of challenge, a frozen vial of D420 was diluted in intravenous-grade saline to the intended challenge inoculum dose. Within one hour from dose preparation, clinical staff administered 0.1 mL of drops containing challenge inoculum in each nostril and participants remained in a semi-supine position for at least 10 minutes.

### Study procedures

#### Participant schedule in the challenge unit

Following informed consent, eligible participants were admitted to CCfV’s challenge unit the day before challenge. All Days (inpatient and outpatient) cited hereafter are defined as the number of days post-challenge. Once participants were inoculated, they completed a standard inpatient observation period of 16 to 24 Days unless shortened by earlier onset of symptoms and initiation of azithromycin treatment. After discharge, participants were monitored as outpatients up to Day 365. The present study reports results to Day 56 when sample testing for *B. pertussis* was completed. Tables S1 and S2 (Appendix A) provide the study schedule for procedures and samples.

#### Azithromycin treatment and discharge criteria

All participants at each dose received a five-day course of azithromycin for infection treatment: 500 mg was provided once on the first day followed by 250 mg once per day for four days.^20^ For participants without evidence of *B. pertussis* infection, the first dose of azithromycin was administered on their discharge day (Day 16) and remaining doses completed as outpatients. Participants positive for *B. pertussis* but not reporting symptoms were started on treatment at the end of the observation period and remained as inpatients until completion. For participants positive for *B. pertussis* with pertussis symptoms, treatment start was determined by timing of symptoms, but no later than Day 19 or 24-48 hours from cough onset, whichever came first. Early treatment in the case of reported mild cough was designed to prevent progression to a paroxysmal cough.^6^ Infected participants were discharged from the challenge unit once treatment was completed and/or showed evidence of negative *B. pertussis* PCRs and an improved clinical assessment, when applicable.

#### Adverse events

Adverse events (AEs) were monitored daily during the inpatient period. Diary cards for recording AEs were distributed with instructions on discharge day and then reviewed by study staff during outpatient visits until Day 42. AEs were assessed for severity as: Grade 1 (mild, not interfering with everyday activities); Grade 2 (moderate, interfering with everyday activities but does not require medical treatment); Grade 3 (severe, preventing everyday activities and requires medical treatment); or Grade 4 (potentially life threatening). Serious adverse events (SAEs) included any event that was life-threatening, caused death, required inpatient hospitalization, caused significant disability or incapacity, or was judged to be serious by the Principal Investigator.

Adverse events of special interest (AESI) were defined as any untoward event determined to be related to the challenge. AESI included AEs that arose from intranasal administration of the challenge inoculum or any clinically severe pertussis disease symptoms (≥ Grade 3) or outcomes (epistaxis, aspiration (evidence of aspiration of the challenge inoculum), paroxysmal cough, post-tussive emesis, and inspiratory whoop, pneumonia, otitis media, and/or cough for more than 3 weeks).

#### Testing for B. pertussis and immunology assay

Nasal washes were collected from each participant at baseline, daily during the inpatient period, and on outpatient Days 21, 28, 35, 42, and 56, for pertussis culture and PCR testing. For nasal washes, a 5 mL normal saline wash of each naris was used to recover mucosal secretions. Culture testing of nasal wash was conducted on charcoal blood agar plates. For PCRs, nasal washes were evaluated by a quantitative PCR assay that assessed amplification of the *B. pertussis* marker IS*481*.

Blood samples for ELISAs to assess anti-PT, -FHA, -FIM, and -PRN IgG were collected at baseline, inpatient days, and outpatient Days up to Day 56 post-challenge (table S2). Seroconversion for each antibody was determined as follows. With baseline titres of less than, or equal to, the specific antigen’s lower limit of detection (LLOD), seroconversion was defined as a ≥4-fold increase up to Day 56. With baseline titres greater than the LLOD, seroconversion was defined as a ≥2-fold increase up to Day 56. Results are reported in international units (IU/mL), except for anti-FIM IgG (EU/mL). Further information on laboratory procedures for cultures, PCRs, and ELISAs are provided in Appendix A.

#### Pertussis symptom assessments

Symptoms of early pertussis disease reported by participants were captured by clinical staff daily during the inpatient period and via participant Diary Cards during the outpatient period up to Day 42. Three solicited systemic symptoms were recorded:

- fatigue
- malaise
- fever

There were seven solicited respiratory systems:

- nasal congestion
- runny nose (rhinorrhea)
- sneezing
- sore throat
- watery eyes
- red eyes
- cough

While all ten symptoms were studied, development of a cough was of specific focus due to its particular association with disease complications.^3,6^ Only when a symptom was reported as Grade 3 or 4 was it captured as an AE.

### Assigning pertussis clinical outcomes

An adjudication analysis by five board certified infectious disease clinicians was used to develop a clinical definition algorithm. This algorithm applies standardized criteria that balances detection of infection from culture and PCR plus symptom association with pertussis to assign clinical outcomes. Using this algorithm, each participant was assigned to one of three possible pertussis clinical outcomes:

> ***Non-Infected —*** Absence of positive culture and less than three positive PCR results for *B. pertussis* on or after Day 6 post-challenge up to first day of azithromycin treatment *regardless* of reporting solicited symptoms. When symptoms were reported during this time frame, but in the absence of confirmed *B. pertussis* detection with daily testing, they were characterized as background symptoms, not pertussis-related symptoms.
>
> ***Symptomatic Infection*** — At least one positive culture and/or three or more positive PCR results for *B. pertussis* on or after Day 6 post-challenge to the first day of azithromycin treatment *plus* a pertussis-related symptom profile during infection, defined as two or more solicited pertussis symptoms (Grade 1 or 2), at least one of which must be a respiratory symptom.
>
> ***Asymptomatic Infection*** *—* At least one positive culture and/or three or more positive PCR results for *B. pertussis* on or after Day 6 post-challenge up to first day of azithromycin treatment *without* a pertussis-related symptom profile.

Assessment of infection status using culture and PCR began on Day 6 post-challenge to eliminate potential misinterpretation of residual challenge inoculum in nasal wash as infection. Test results for *B. pertussis* in nasal wash (colony count or PCR) were always assessed prior to evaluating symptoms.

Overall infection rates included asymptomatic plus symptomatic infection outcomes. Symptomatic infections with and without a cough were also assessed and recorded.

### Statistical Analysis

Descriptive statistics were used to summarize pertussis outcome frequency per dose group, study enrollment, demographics, vaccine priming status, bacterial load, symptoms, bacterial clearance, and antibody responses among participants. Safety data including AEs, SAEs, and AESI were tabulated and summarized. An exploratory generalized linear mixed model (GLMM) was used to model the probability of reporting each solicited symptom, with days post-challenge, clinical outcome, and their interaction included as fixed effects, and participant included as a random effect to account for repeated observations. Due to the study being exploratory, we do not present p-values for our analyses as there were no *a priori* hypotheses. No imputation was performed for missing data. Analyses were conducted using all available data points. All statistical analyses and tabulations were performed in SAS and R.^21,22^

## Results

Among 168 screened individuals, 79 eligible participants were consented to the study and challenged with a dose of *B. pertussis* (CONSORT diagram, figure 1). Of those, 75 participants were included in analysis (figure 1). Four participants were excluded from analysis; three withdrew during the inpatient period (unrelated to *B. pertussis* exposure) and one developed a confounding *Bordetella bronchiseptica* infection after screening, whose positive baseline (Day -1) results became available after challenge (figure 1). A summary of participant disposition is provided in table S3. Demographics of all challenged participants under study, and the subset in analysis of the four highest doses (stage two), are shown in table 1. All participants had been immunized in infancy with an aP (33·3%) or wP (66·6%) vaccine (table S4). Doses administered ranged from 10^4^ CFU to 10^8^ CFU (figure 1).

**Table 1.** Demographic characteristics of all 75 challenged participants under study, and for the subset included in the stage-two analysis.

| Characteristic | Categories | All doses<br>(n=75) |  | Stage-two doses<br>(n=53) |  |
| --- | --- | --- | --- | --- | --- |
|  |  | n | % | n | % |
| Age at enrollment (years) | Median | 29 | .. | 26 | .. |
|  | Min | 19 | .. | 19 | .. |
|  | Max | 40 | .. | 40 | .. |
| Sex assigned at birth | Male | 39 | 52 | 30 | 56.6 |
|  | Female | 36 | 48 | 23 | 43.4 |
| Self-described ethnicity or race* | White | 63 | 84 | 43 | 81.1 |
|  | Black | 3 | 4 | 3 | 5.7 |
|  | Chinese | 3 | 4 | 3 | 5.7 |
|  | Filipino | 4 | 5.3 | 3 | 5.7 |
|  | Arab | 1 | 1.3 | 1 | 1.9 |
|  | Latin American | 1 | 1.3 | 1 | 1.9 |
|  | South Asian | 1 | 1.3 | 0 | 0 |

\*Participants could select multiple categories for ethnicity or race
n = number of participants
%= percentage of participants
Additional demographic data are provided at Clinicaltrials.gov (NCT05136599)

### Symptomatic infection endpoint

#### Stage one: Identification of the HID70-90

Starting at 10^4^ CFU of D420 (figure 1), the first dose that led to preliminary identification of HID70-90 was 10^7^ CFU (table 2). Following the study plan, 6 additional 10^7^ CFU challenges were then administered for a total of 12 participants at this dose. Overall, a symptomatic infection rate of 75·0% (9 of 12 participants) established the HID70-90 at 10^7^ CFU, allowing the study to progress to stage two.

**Table 2.**
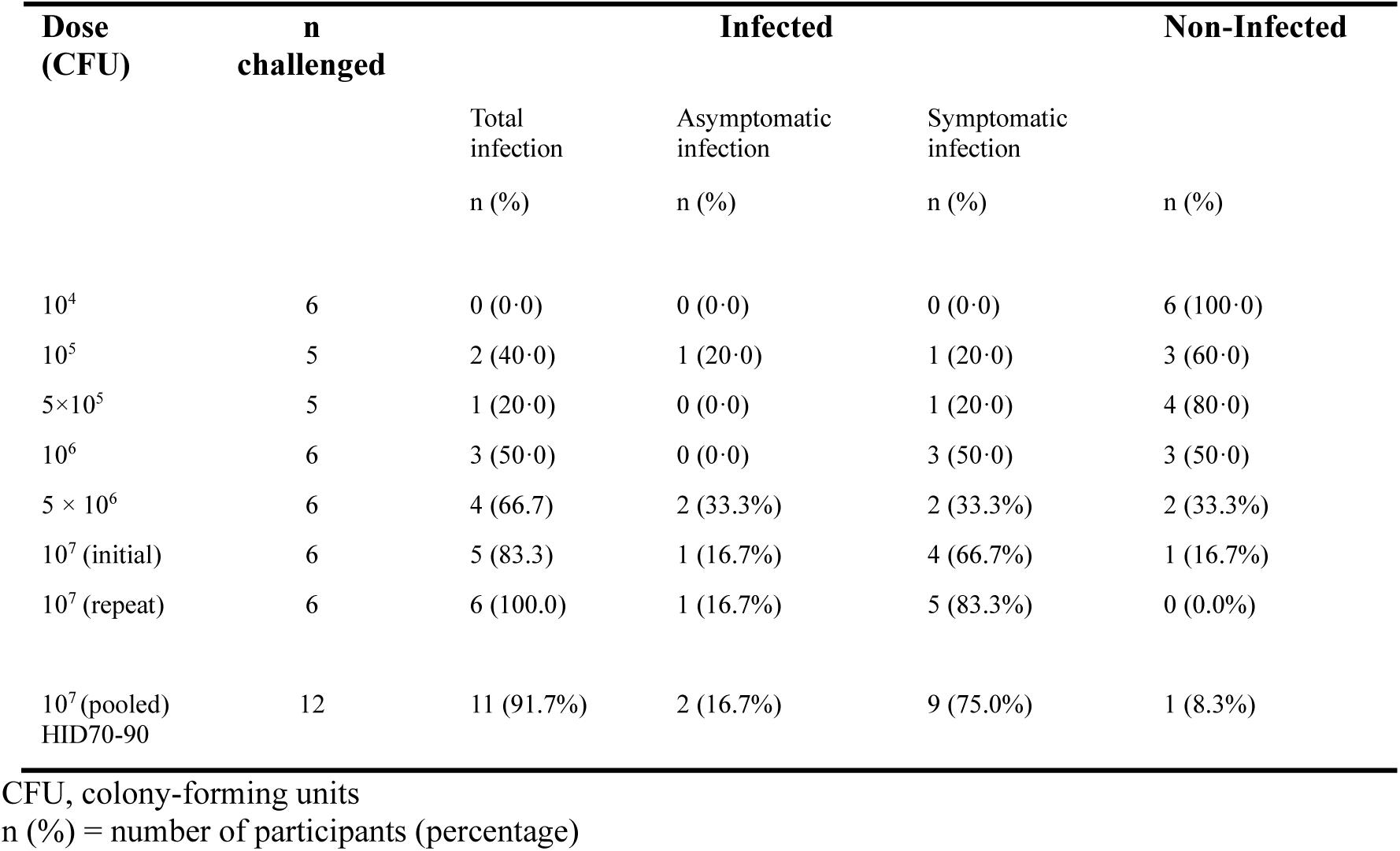
Participant clinical outcomes for the dose escalation in stage one.

#### Assessments of stage-two doses

The four doses investigated in stage two were 5×10^6^, 10^7^, 5×10^7^, and 10^8^ CFU (n=53, table 3). These doses induced the highest rates of overall infection (symptomatic plus asymptomatic) among all studied doses (tables 2 and 3). Overall infection rates observed were 60·0%, 86·4%, 91·7%, and 88·9%, respectively. Symptomatic infection rates were 20·0%, 72·7%, 58·3%, and 55·6%. With these additional challenges, the HID70-90 was further verified at 10^7^ CFU (table 3).

**Table 3.** Participant clinical outcomes for the four uppermost doses in the stage-two analysis.

| Dose (CFU) | n stage-two analysis* | Infected |  |  | Non-Infected |
| --- | --- | --- | --- | --- | --- |
|  |  | Total infection | Asymptomatic infection | Symptomatic infection |  |
|  |  | n (%) | n (%) | n (%) | n (%) |
| 5×10<sup>6</sup> | 10 (6+4) | 6 (60·0) | 4 (40·0) | 2 (20·0) | 4 (40·0) |
| 10<sup>7</sup> (HID70-90) | 22 (12+10) | 19 (86·4) | 3 (13·6) | 16 (72·7) | 3 (13·6) |
| 5×10<sup>7</sup> | 12 | 11 (91·7) | 4 (33·3) | 7 (58·3) | 1 (8·3) |
| 10<sup>8</sup> | 9 | 8 (88·9) | 3 (33·3) | 5 (55·6) | 1 (11·1) |

CFU, colony-forming units
n (%) = number of participants (percentage)
\*For 5×10<sup>6</sup> CFU and 10<sup>7</sup> CFU, the values in parenthesis indicate the number of challenged participants from the dose escalation (stage one) plus and those added in the stage-two challenges respectively. For 5×10<sup>7</sup> and 10<sup>8</sup> CFU, all challenges were conducted in stage two. Study-wide n values pooled across stage one and stage two were 35, 15, 25 for symptomatic infection, asymptomatic infection, and non-infected, respectively.

#### Dose accuracy

The doses prepared for participant challenges were highly accurate and closely matched the targeted dose, as shown in table S5. All doses prepared for challenges were within three times the intended dose. Instances of a prepared dose overlapping with another target dose only occurred when two dose groups differed by two-fold. No cases of a prepared dose overlapping with another dose group were observed wherever target doses differed by five- or ten-fold.

### Symptoms related to pertussis infection

All solicited symptoms were reported among individuals who showed evidence of infection (symptomatic infection) as well as those who did not (non-infected), with the exception of fever that was non-reported in both groups (0%). Using GLMM analyses, we evaluated the relationship between each of the nine reported symptoms and pertussis infection, accounting for background symptoms. In the models, non-infected participants provided a proxy for background non-specific symptoms. Study-wide analysis was conducted to improve sample size values per group given that non-infected rates decreased at higher doses. Overall, each of the solicited symptoms reported in the symptomatic infection group (n=35) was more common with time post-challenge than those in the non-infected group (n=25), except for sore throat which became more common in both groups (figure S1A-I). The average time to the onset of solicited symptoms with infection was 10–12 days post-challenge.

### Characterization of a symptomatic CHIM

Once the HID70-90 was established, the four highest doses were used to thoroughly characterize the emerging pertussis model. Post-challenge load of *B. pertussis* in nasal wash samples, pertussis symptoms, and antibody responses were each evaluated over time. Geometric mean culture colony counts and mean cycle threshold (Ct) for PCRs of the *B. pertussis* marker IS*481* demonstrated that the bacterial load in nasal wash increased with the challenge dose (figure 2A, B; confidence intervals (CI) in S2A, B). Increased bacterial load in nasal wash was also observed as infection progressed from asymptomatic, to symptomatic, and finally to symptomatic with cough (figure 2C, D, figure S2C, D).

**Figure 2.**
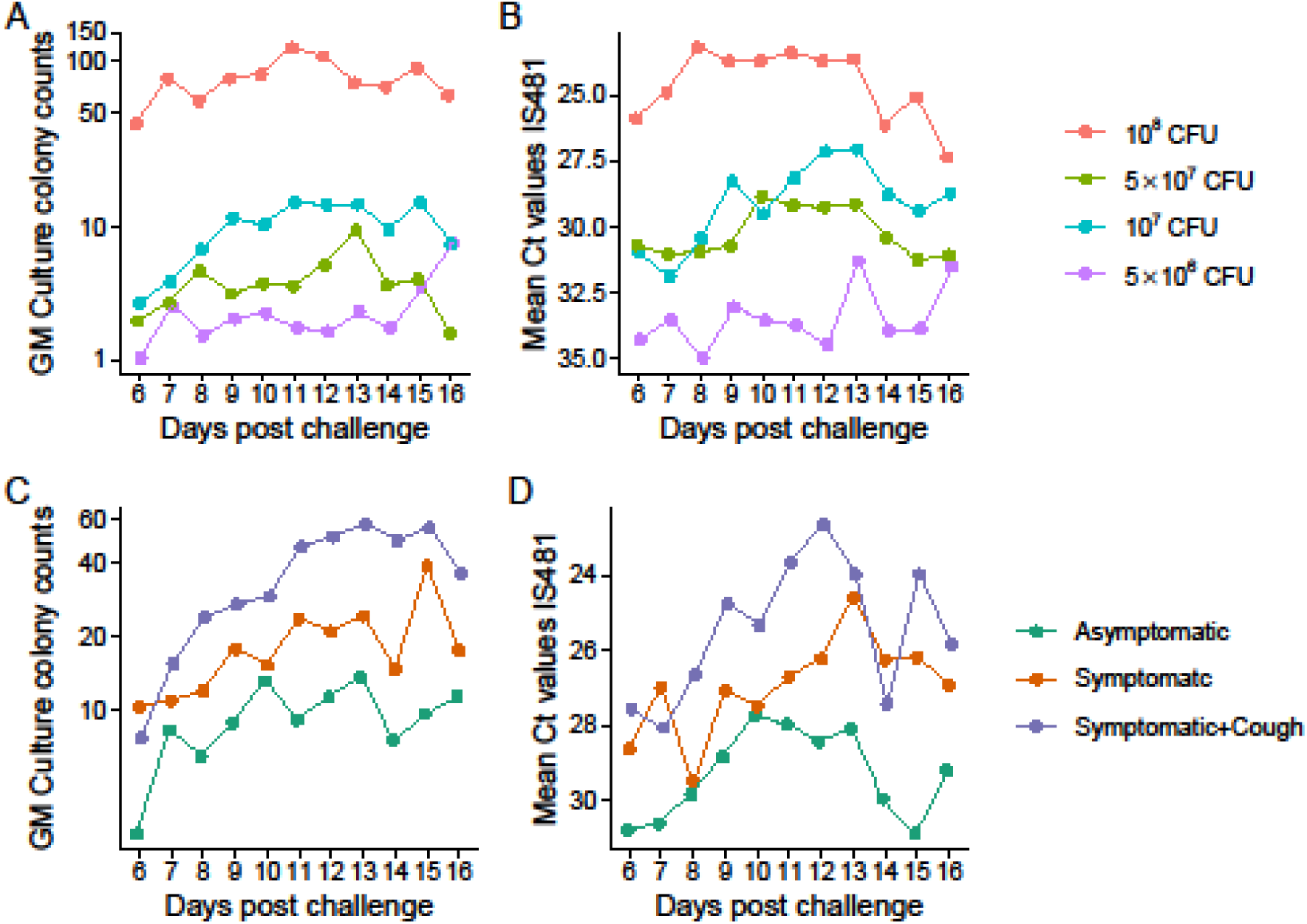
*B. pertussis* detection (average) in nasal wash over time post-challenge for the stage-two analysis of the four highest studied doses. A) geometric mean (GM) colony counts by days; B) mean cyle threshold (Ct) IS*481* values by days; C) GM colony counts by asymptomatic infection, symptomatic infection, and symptomatic infection with cough and; D) mean Ct IS*481*values by asymptomatic infection, symptomatic infection, and symptomatic infection with cough. Confidence intervals are in figure S2.

Symptoms that were at least two-fold more common among participants assigned symptomatic infection (n=30) than non-infected (n=9) for the four highest doses were nasal congestion, fatigue (nearly seven-times more common), malaise, runny nose, and cough (figure 3). Sneezing was marginally more frequent for symptomatic infection (by 1.4-fold). Sore throat had similar frequency between groups. Ocular symptoms were the least common overall. Cough was reported for 50.0% of symptomatic infection cases at the four highest doses (figure 3, table S6), and also for the HID70-90 of 10^7^ CFU alone (figure S3A; for symptoms across all doses, figure S3B). Fever was non-reported at the four highest doses (and study-wide (0%)).

**Figure 3.**
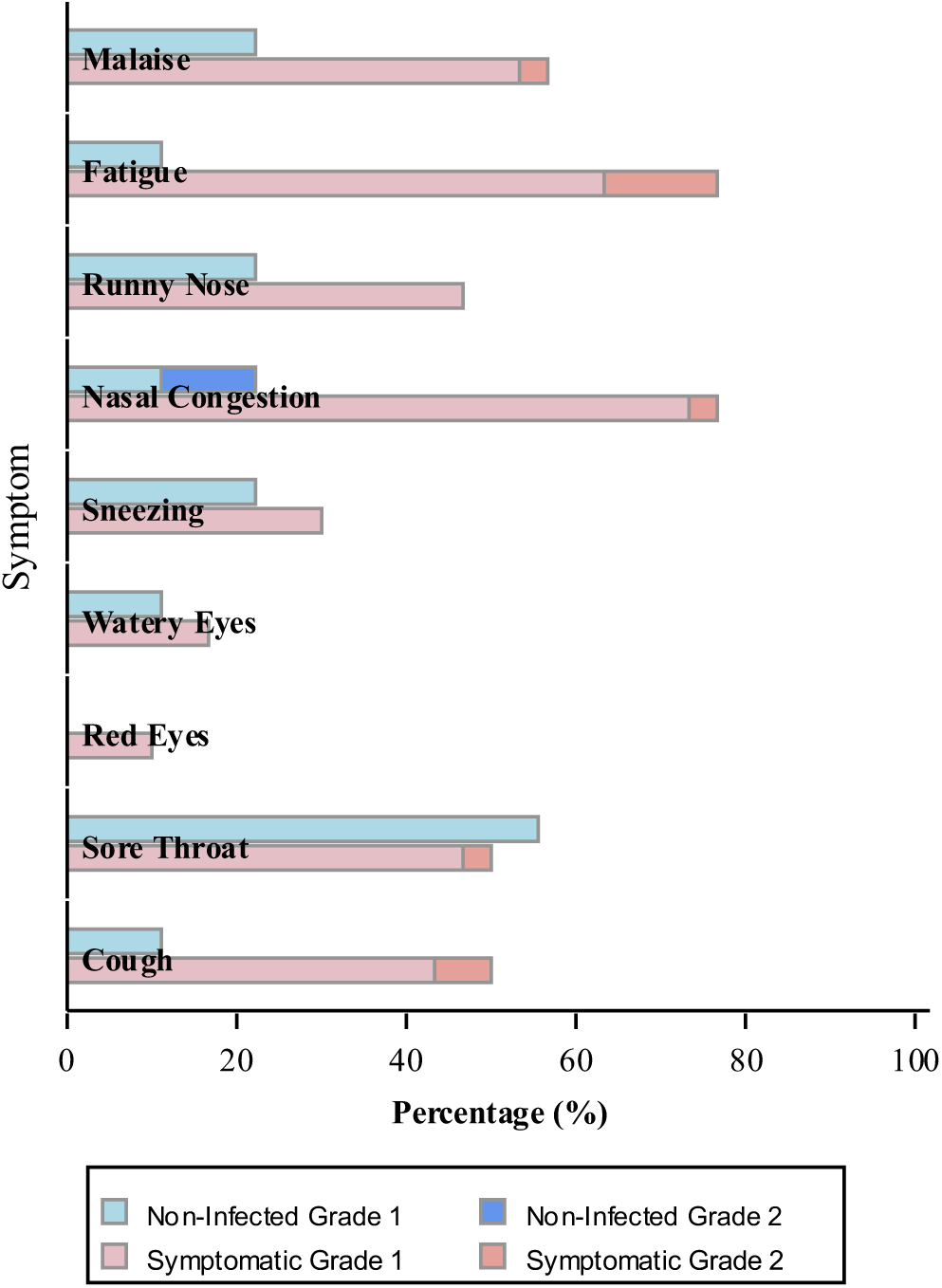
Percentage of participants with solicited symptoms (all Grade 1 or 2) for non-infected and symptomatic infection across the four uppermost doses (stage two)

Geometric mean titres (GMTs) of anti-PT, -FHA, -FIM, and -PRN IgG to Day 56 for the four highest challenge doses are shown in figure 4 (CIs in figure S4). GMTs for these antibodies increased with time post-challenge for each dose group, with generally higher titres with increasing challenge dose. GMTs by clinical outcome revealed elevated titres for symptomatic infection as compared to asymptomatic infection and non-infected for each antibody, except anti-PRN IgG that had similar GMTs for symptomatic and asymptomatic infection (each higher than non-infected, figure S5). Seroconversion for at least one of the four studied antibodies was observed for 35.7% of participants with asymptomatic infection and 70.0% of those with symptomatic infection. None of the non-infected participants seroconverted (table S7; for seroconversion rates across all doses, table S8).

**Figure 4.**
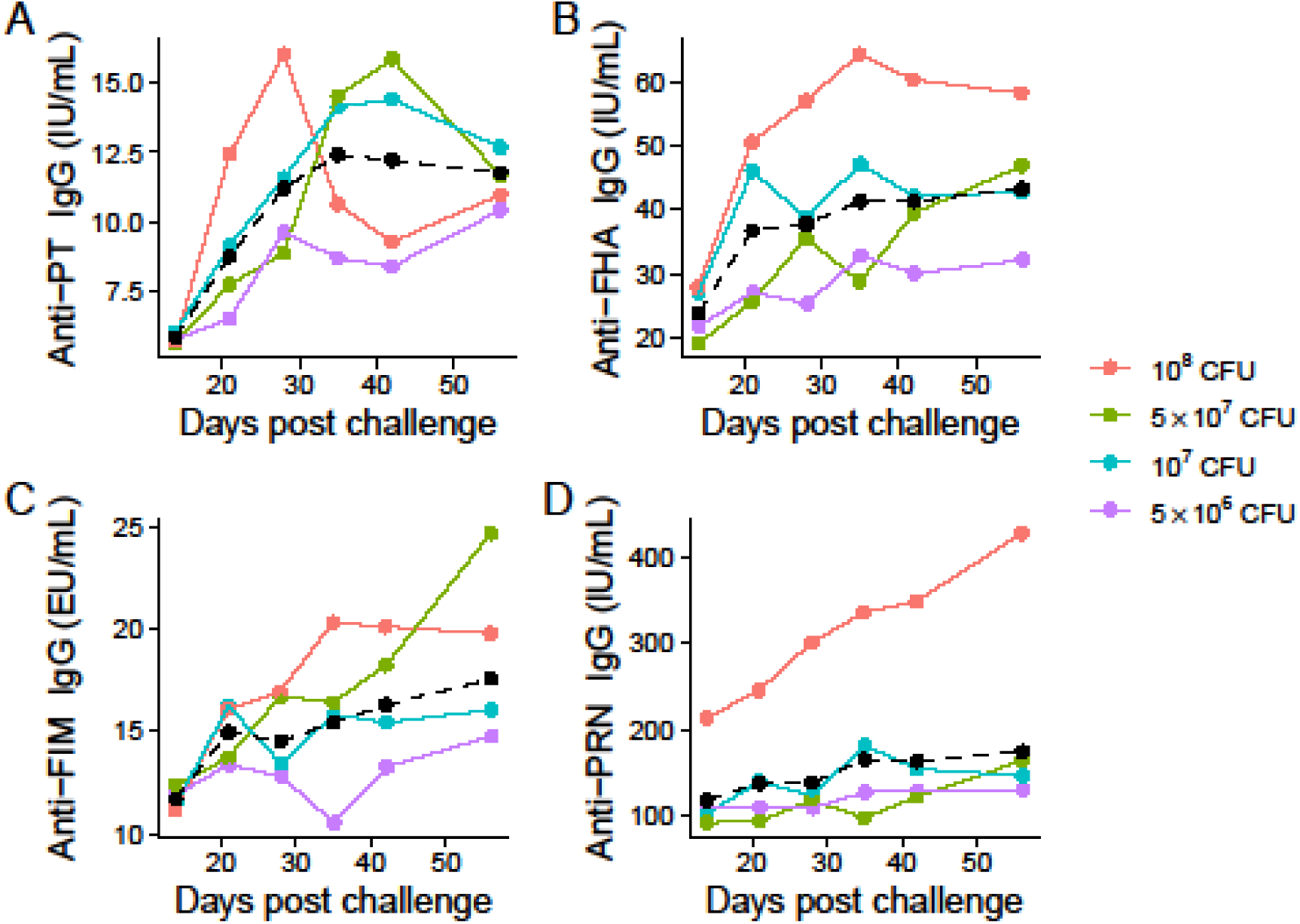
Geometric mean titres over time post-challenge for pertussis antibodies at the four uppermost (stage-two) doses. A) anti-PT IgG; B) anti-FHA IgG; C) anti-FIM IgG and; D) anti-PRN IgG. The dotted line indicates the average among groups. Confidence intervals are in figure S4.

### Vaccine priming

Participants primed with aP vaccination in infancy exhibited higher *B. pertussis* load in cultures (greater colony counts) and PCRs (lower Ct values) as compared to wP primed participants (figure S6A, B). The elevated *B. pertussis* load in those who were aP primed as compared to wP primed was consistently observed across all studied inpatient days.

### Adverse Events

Collectively, 59 (74·7%) of the 79 challenged (exposed) participants reported one or more unsolicited AE (table S9). Of these, there were three study-related AEs (3·8%): two cases of prolonged bacterial shedding arising post-treatment during the outpatient period (Grade 1), which were successfully detected and managed accordingly, and one case of a Grade 1 swollen lymph node.

Seven participants (8·9%) had ≥1 AE classified as ≥Grade 3 in severity, all of which were unrelated to *B. pertussis* challenge administration or infection. One Grade 3 AE was anxiety related to challenge-room isolation and led to the participant’s withdrawal during their inpatient stay. No AESI were reported. Three SAEs were recorded for one participant during the outpatient period due to a motor vehicle accident and were unrelated to the study. There were no study-related SAEs. No pertussis symptoms were reported as Grade 3 or 4.

### Azithromycin treatment

All infected participants cleared *B. pertussis* as indicated from nasal samples by the end of the five-day treatment with azithromycin (figure S7A, B, table S10). For symptomatic infection, the proportion of participants with symptoms (including cough, figure S8) decreased during and shortly after the five-day azithromycin course and all symptoms were non-persistent.

## Discussion

In this study, we established a pertussis CHIM that induces early symptomatic pertussis disease with no challenge-related safety events. A 10^7^ CFU *B. pertussis* D420 challenge dose was the lowest, and only dose, identified as the HID70-90, with approximately 73% of all challenged participants exposed developing symptomatic infection, half of which reported cough. The four highest challenge doses evaluated (5×10^6^, 10^7^, 5×10^7^ and 10^8^ CFU) provided insights into the rates of infection, bacterial load, symptom presentation, and antibody responses, that further affirm a safe and reliable pertussis CHIM.

### CHIM characterization

To our knowledge, a symptomatic pertussis CHIM for a wild-type strain has not been reported previously.^13^ Our study is proof-of-concept for a safe symptomatic pertussis CHIM established by applying a rigorous study design that includes reliable dose accuracy supporting incremental dose increases, effective rescue therapy, and close inpatient and outpatient monitoring of participants’ infection and symptoms with careful assessments by the DSMB. We affirmed the symptomatic endpoint as evidenced by an elevated frequency of solicited low-grade symptoms for those with confirmed infection (compared to non-infected), particularly for nasal congestion, runny nose, fatigue, malaise, and cough, emulating adult pertussis manifestation in real-life settings. The lack of fever concurs with reports that this symptom is absent or rare for pertussis.^14,23^ Overall, the study provides a pathway for additional symptomatic CHIMs for pertussis.

#### Controlling for background symptoms

While our study did not include a control group *sensu stricto* (placebo), symptoms reported by the non-infected group served to distinguish background symptoms from pertussis-related symptoms for the infected group. The alignment between increased symptom frequency (relative to background symptoms for non-infected) and the timing of their onset at ten or more days post-challenge in the symptomatic infection group, which corresponds to the natural pertussis incubation period,^6^ supports the conclusion that symptoms were triggered by the challenge and were infection-related.

#### Effects of challenge dose

The finding that higher D420 challenge doses were associated with greater *B. pertussis* load in nasal wash, and particularly for symptomatic infection with cough, demonstrates a dose-response relationship. Further studies using this CHIM should investigate the relationship between inoculum dose and cough frequency and severity (Grade 1 or 2) to elucidate physiological mechanisms. Such research may help ascertain whether a cough, which often underlies pertussis disease complications,^2,3,23^ is driven by bacterial load.

Among the four highest doses studied, the symptomatic HID70-90 was observed only at 10^7^ CFU (∼73%). The overall infection rates at the two highest doses, 91·7% and 88·9% for 5×10^7^ and 10^8^ CFU respectively, were similar to, or slightly higher than, that observed at 10^7^ CFU (86·4%). Notably, however, this did not translate into increased symptomatic infection rates at those two uppermost doses (58.3% and 55.6%, respectively). Differences in symptom onset and clinical expression post-infection may reflect heterogeneity in immune histories, as none of the participants were vaccine-naïve and prior natural infection could not be fully ruled out. Variability in the onset of disease concurs with community-based research. A meta-analysis of laboratory-confirmed pertussis cases demonstrated that only a subset of infected household contacts developed symptomatic disease, with 5% to 56% remaining asymptomatic.^24^ Collectively, interindividual host-pathogen dynamics, shaped by prior immunologic exposure, host genetics, microbiome,^25^ and other factors, may contribute to variability of symptomatic infection rates in this CHIM, mirroring patterns observed at the population level.

#### Antibody responses

This CHIM reproducibly elicited an antibody response as demonstrated by rising titres of all four evaluated antibodies (anti-PT, -FHA, -FIM, and -PRN IgG) assessed through to Day 56 across the four uppermost inoculum doses. Symptomatic infection was associated with boosted titres as compared to asymptomatic infection for anti-PT, -FHA, and -FIM IgG, while anti-PRN IgG titres were similarly boosted by both symptomatic and asymptomatic infection. Enhanced production of these four antibodies in response to vaccination (aP or wP) or natural infection has been associated with protection against pertussis disease (that wanes over time).^1,5,26,27^ The current CHIM provides a platform to further study these antibody pathways and correlates of disease protection.

Seroconversion occurred exclusively among participants classified as infected (asymptomatic or symptomatic) and was not observed for any non-infected. Participants with symptomatic infection exhibited approximately a two-fold higher rate of seroconversion for at least one antibody compared to those with asymptomatic infection at the four highest doses, that is consistent with augmentation of antibody responses with clinical disease. We also noted that not every infection led to seroconversion which may be due, in part, to early initiation of antimicrobial treatment according to protocol. Halting infection progression and clearing bacterial antigens may attenuate antibody responses.^8^

Overall, the serological findings concurred with the pertussis outcomes assigned by the clinical definition algorithm using infection and symptom measures (non-infected, asymptomatic infection, symptomatic infection). No seroconversion was observed in participants assigned as non-infected by the algorithm. This approach offers a standardized tool to assess infections in future studies, minimizing potential variability introduced by investigator-dependent assessments.

#### Symptomatic and asymptomatic infections are safe

We demonstrated that all doses evaluated between 10^4^ to 10^8^ CFU were safe and did not result in any severe symptoms, including cough. Two cases of prolonged *B, pertussis* shedding and one swollen lymph node (all Grade 1) related to challenge were detected due to rigorous clinical assessments and outpatient testing and were effectively treated or managed. In sum, the fact that there were no study-related SAEs, and that azithromycin cleared all infections and diminished symptoms, including cough, supports the safety of this CHIM for healthy adult participants.

### Vaccine priming status

A strength of this study is the inclusion of participants with either aP or wP vaccination in infancy.^8^ This provides an opportunity to explore how early life vaccine priming may be associated with susceptibility to infection. We found that the *B. pertussis* load in nasal wash was greater for participants primed with aP-than wP over time post-challenge. We also showed an increased *B. pertussis* load in individuals classified as having symptomatic infection (with and without a cough) than for asymptomatic infection. Together, these results suggest that aP vaccine priming may be less protective than wP against infection and progression to symptomatic disease, paralleling findings from experimental studies in baboons and population studies in humans.^1,4,5,28^ Future research with additional participants using this CHIM will allow stratification of priming status by demographics, antibody responses, and rates of symptomatic infection.

### Study limitations

This study should be interpreted in the context of certain limitations. First, the sample sizes (n=5 to 6 participants) during dose-escalation stage to safely identify the HID 70-90 were small by design, and statistical variability of the estimated percentages for clinical outcomes were elevated at these smaller sample sizes. Second, similar to other CHIMs, healthy adults were studied and thus aspects of our results may not be generalizable to young children, elderly, or immunocompromised individuals, who are most susceptible to pertussis complications.^3^ Third, the trial occurred in a controlled, and participant-isolated environment, without potential for inter-person transmission and thus may not parallel all aspects of naturally acquired disease. Significantly, the presence of early pertussis respiratory symptoms and elevated bacterial load with greater challenge dose in this CHIM may provide opportunities to study *B. pertussis* infection transmission.^29^ Finally, the results for the pertactin-producing isolate D420 may differ from other pertussis isolates. D420 represents a previously predominating strain in North America, from a clade of wild-type isolates harboring *ptxP3, prn2* and *fimH2 (fim3-2)*^17–19^ that was isolated in the USA in 2002.^12,19^ Since then, it has been used as a virulent isolate for experimental pertussis research in animal models.^12,18^ This study provides a foundation for development of additional CHIMs using strains of current, or potential future, epidemiological threat including pertactin-deficient strains currently circulating in North America.^18,30^

### Conclusions

Our study demonstrates the establishment of a safe and reproducible symptomatic pertussis CHIM using *B. pertussis* D420. The CHIM provides opportunities to study clinical, microbial, immunological, and pathological aspects of pertussis infection and disease. By titrating the challenge dose, novel vaccines and therapeutics may be rapidly assessed against a spectrum of infection endpoints including symptomatic-, asymptomatic- and non-infection.

## Data Availability

All data produced are available online at Clinicaltrials.gov (NCT05136599)

## Contributors

**MSE:** conceptualization, data curation, funding acquisition, investigation, methodology, project administration, resources, supervision, validation, writing–original draft. **KLR:** conceptualization, data curation, investigation, methodology, project administration, resources, validation, writing–original draft. **JML:** conceptualization, investigation, methodology, validation. **LY:** data curation, formal analysis, software. **WB:** data curation, formal analysis, software, visualization. **BS:** conceptualization. **JW:** conceptualization, methodology. **BA:** investigation, methodology, validation. **JHF:** conceptualization, resources, **KME:** conceptualization, methodology, validation. **CBC:** conceptualization. **SM:** conceptualization, investigation. **TFH:** conceptualization, investigation. **JJL:** conceptualization, methodology, resources. **SH:** conceptualization, supervision. **LP:** conceptualization, data curation, methodology, supervision. **PM:** conceptualization, data curation, methodology. **LMF:** conceptualization, data curation. **CAW:** methodology, validation, visualization, writing–original draft. **SAH:** conceptualization, data curation, funding acquisition, investigation, methodology, resources, supervision, validation, writing–original draft. All authors contributed towards the review and editing of the manuscript and agreed with the decision to submit for publication.

## Data sharing

The data are publicly available at Clinicaltrials.gov (NCT05136599).

## Acknowledgements

The authors sincerely thank all participants for their time and commitment to this study. We are grateful to the Challenge Material Production Team, Clinical Staff, Laboratory Staff, and Data Management & Analysis Staff, including Project Managers and Administrators, at the Canadian Center for Vaccinology for their diligence and dedication to the success of this project.

## Supplementary Material

**Appendix A.** Extended Methods, Supplementary Tables and Figures.

