## Appendix A for "A controlled human infection model for symptomatic pertussis in North America using the pertactin-producing clinical isolate D420"

ElSherif et al.

### Supplementary Methods

#### Challenge administration

Every challenge dose prepared for a participant was enumerated and tested for purity and accuracy.

#### Study procedures

Nasopharyngeal aspirates were collected in addition to nasal washes (Table S2), and the latter were used for *B. pertussis* level analysis, which from prior study are most sensitive.[1]

For culture, charcoal blood agar plates supplemented with 0.04 g/L cephalexin were inoculated with 100 µl of neat NW sample and incubated at 36°C for 3 – 7 days. No growth at 7 days was reported as "negative". For PCR, 400 µl of vortexed nasal washes were added to extraction buffer tubes (442817 BD Company, Sparks, Maryland, USA) and placed in system racks containing real-time multiplex pertussis PCR assay reagents (450-003-C-MAX, by BioGX, Birmingham, Alabama, USA); BD MAX operation steps were followed as per manufacturer's instructions. Exponential amplification of IS*481* was considered "positive" regardless of cycle number (under 40 cycles).[2]

Pertussis serum antibodies were measured using a validated enzyme-linked immunosorbent assay (ELISA) with reference sera standardized against U.S. Reference Pertussis Antiserum (Human) Lot 3, 4 or WHO International Standard 06/140. Pertussis antigens were adsorbed onto the wells of labelled polystyrene flat-bottomed plates (Nunc Polysorp, Roskilde, Denmark) at different concentrations (PT from List Biological Laboratories, Inc., Campbell, CA, USA: 0.5µg/mL; FHA from Connaught: 0.5µg/mL; PRN from Sanofi: 1.5µg/mL; FIM from Sanofi: 0.75µg/mL) and incubated for 16-24 hours. This and all assay incubations were in a rotator/incubator set at 28ºC/100 RPM. Plates were washed 3 times between steps with saline (in house) and 0.05% Tween-20 (Sigma). Reference sera, control sera and test sera were serially diluted in incubation buffer (in house), then 100µL or 50 µL added to the antigen coated wells and incubated for 2.5 hours. Next, 100µL of phosphatase-labeled goat anti-human IgG antibody (KPL, Gaithersburg, MA, USA) were added to all wells and incubated for 16-24 hours. Lastly, 100µL of 1 mg/mL p-nitrophenyl phosphate (Sigma, city country) in substrate buffer (1M Tris/.3mM MgCl*2*, pH 9.8) was added to the wells, and after 60 minutes absorbance per well was measured at 405nm using a Biotek Cytation 1 Imaging Reader. Total serum levels of IgG against PT, FHA, PRN and FIM were computed using a reference line calculation software (Gen5 Software version 3.11) and reported as international units per milliliter (IU/mL) for all antigens, except FIM which was in ELISA units per milliliter (EU/mL); the lower limits of quantification (LLQs), were 10 IU, 10 IU, 10 IU, and 16 EU, respectively.

#### Eligibility criteria

Inclusion criteria

To be eligible for the study, each participant must have satisfied ALL of the following criteria:

1. Age 18–40 years, inclusive
2. Good general health status, as determined by history and physical examination conducted no longer than 30 days prior to the challenge
3. Participants who, in the opinion of the Investigator, can and will comply with the requirements of the protocol (e.g., complete Diary Cards, return for follow-up visits).
4. Written informed consent obtained from the participant.
5. If female of childbearing potential and heterosexually active, has practiced adequate contraception for 28 days prior to challenge and has a negative pregnancy test on the day before *B. pertussis* challenge and agreed to continue adequate

contraception until 60 days after inoculation. Adequate contraception is defined as a contraceptive method with a failure rate of <1% per year when used consistently and correctly and, when applicable, in accordance with the product label.

6. Fully vaccinated against SARS-CoV-2/COVID-19 according to provincial Public Health guidelines.
7. If there was a reported history of SARS-CoV-2/COVID-19 infection, the participant must be asymptomatic for >4 weeks.

Exclusion criteria

Participants meeting any of the following criteria were excluded:

1. Underlying chronic medical condition requiring ongoing follow-up and monitoring by a physician (e.g., diabetes, seizure disorder).
2. Underlying cardiac and/or pulmonary disease including hypertension, angina, prior myocardial infarction, asthma, emphysema, chronic bronchitis, and pulmonary tuberculosis.
3. Moderate or severe symptoms of health anxiety, anxiety, and mood symptoms. Self-reported current diagnosis of a major psychiatric illness, including a schizophrenia spectrum disorder, bipolar disorder, posttraumatic stress disorder, obsessive compulsive disorder, substance use, or eating disorder.
4. QT prolongation on electrocardiogram (EKG).
5. History of everyday smoking/vaping in the last 2 years and/or current smoking/vaping more than once per week.
6. Pregnant (known before or established at the time of screening using a urine-based test) or breastfeeding.
7. Immunocompromised (with HIV/AIDS-positive or receiving immunosuppressive therapy involving steroids) or with any medical condition or medication that leads to a compromised immune system.
8. Positive for hepatitis B or C.
9. Vaccinated against pertussis within previous 5 years and/or >7 cumulative doses from infancy to date of screening.
10. Reported history of laboratory-confirmed pertussis infection
11. Antibody titre to pertussis toxin >20 IU/mL (2X the lower limit of quantification (LLOQ).
12. Nasopharyngeal detection of *B. pertussis* prior to challenge using culture isolation and/or PCR detection, or detection of other respiratory infection.
13. Living with young children (<1 year of age) or with any household member not current in their pertussis immunization up to Day 56 post-challenge. Note: Household members whose vaccination is not current were offered a pertussis-containing vaccine, as recommended and funded by the Nova Scotia Department of Health and Wellness.
14. Living or working with (any form of close contact) any of the at-risk/vulnerable groups (children <1 year of age, pregnant woman who have not yet received their maternal Tdap vaccine, immunocompromised individuals, anyone not current in their pertussis immunization, or adults >65 years of age who have not received a dose of Tdap vaccine within the past 10 years) up to Day 56 post-challenge.
15. Known allergy to macrolides including azithromycin or erythromycin, history of Clostridium difficile within last 2 months.
16. Any contraindication to receiving azithromycin.
17. Taking any antibiotic currently or within the previous 2 weeks.

18. Currently taking terfenadine, astemizole, theophylline, or cimetidine.
19. Recent (within 6 months) nasal or sinus surgery, recent use of intranasal steroids (4 weeks), or diagnosis with nasal polyps.
20. Receipt of any investigational drug or vaccine (including SARS-CoV-2/COVID-19 vaccine) within 6 months prior to challenge. An investigational vaccine is defined as a vaccine that is still being tested in clinical trials or one that has not yet been authorized for use in Canada for administration by public vaccine programs.
21. Receipt of any authorized vaccines within 2 weeks of being challenged with *B. pertussis* in this study.
22. Current laboratory-confirmed SARS-CoV-2/COVID-19 infection
23. Previous moderate or severe laboratory-confirmed SARS-CoV-2/COVID-19 infection that required hospitalization
24. Head trauma (e.g., fracture of the cribriform plate) within 1 year of screening.
25. Any other finding that the Investigator considers will make the participant unsuitable for the study or unable to comply with the study requirements.
26. Symptoms indicative of acute respiratory illness (such as fever, cough, difficulty breathing) identified during the physical examination done on Day -1 (check-in) or Day 0 before a participant is challenged.
27. History of Bell's Palsy and/or facial paralysis.
28. Receipt of facial cosmetic filler in the past 3 months.

**Statistical analysis**

To assess symptom development, an exploratory analysis was developed using a generalized linear mixed model (GLMM), The GLMM was based on a binomial distribution and a logistic link function to model the probability of a symptom being present by days post challenge (days 6-16) and outcome group (non-infected versus symptomatic infection) with participant as a random effect. We fit this model to the nine individual symptoms. Missing values were not inputted.

**Supplementary Tables and Figures**

**Table S1. Clinical and safety procedures**

| **Clinical/Safety** | **OPt** | **Inpatient/Challenge Unit** | | | | | | | | | | | | | | | | | **Outpatient (OPt)** | | | | | |
|---|---|---|---|---|---|---|---|---|---|---|---|---|---|---|---|---|---|---|---|---|---|---|---|---|
| | SCR | Check-in | Challenge | | | | | | | | | | | | | | | | Phone Calls | | | | | |
| **Visit** | **1** | **2** | **3** | **4** | **5** | **6** | **7** | **8** | **9** | **10** | **11** | **12** | **13** | **14** | **15** | **16** | **17** | **18** | **19** | **20-23** | **24** | **25** | **26** | **27** | **28** |
| **Day:** | **-30** | **-1** | **0** | **1** | **2** | **3** | **4** | **5** | **6** | **7** | **8** | **9** | **10** | **11** | **12** | **13** | **14** | **15** | **16** | **17-20** | **21** | **28** | **35** | **42** | **56** |
| **Clinical** | | | | | | | | | | | | | | | | | | | | | | | | | |
| Informed Consent | X | | | | | | | | | | | | | | | | | | | | | | | | |
| Screening (MH, Demo) | X | | | | | | | | | | | | | | | | | | | | | | | | |
| Medications | X | | X | X | X | X | X | X | X | X | X | X | X | X | X | X | X | X | X | | X | X | X | X | X |
| Physical Examination | | X | X | X | X | X | X | X | X | X | X | X | X | X | X | X | X | X | X | | | | | | |
| History-directed PE | | | | | | | | | | | | | | | | | | | | | X | X | X | X | X |
| Vital Signs | X | X | X | X | X | X | X | X | X | X | X | X | X | X | X | X | X | X | X | | | | | | |
| Phone Calls | | | | | | | | | | | | | | | | | | | | X | | | | | |
| Clinic Visits | | | | | | | | | | | | | | | | | | | | | X | X | X | X | X |
| **Safety** | | | | | | | | | | | | | | | | | | | | | | | | | |
| Urine Pregnancy Test | X | X | | | | | | | | | | | | | | | | | | | | | | | |
| EKG | X | | | | | | | | | | ? | ? | ? | ? | ? | ? | ? | ? | ? | | ? | | | | |
| HIV, HBV, and HCV Screening (5mL blood) | X | | | | | | | | | | | | | | | | | | | | | | | | |
| NPS for SARS-CoV-2 | X | X | | | | | | | | | | | | | | | | | | | | | | | |
| Pertussis serology screening (3 mL) | X | | | | | | | | | | | | | | | | | | | | | | | | |
| Safety Bloods (5-10mL) / **CBC only | X | X | X** | X** | | X | | X | | X | | | X | | | | X | | X | | X** | X** | | | X** |
| Urinalysis | X | X | | | | | | | | | | | | | | | | | | | | | | | |
| Receive DC | | | | | | | | | | | | | | | | | | | X | | | | | | |
| DCs collection/review | | | | | | | | | | | | | | | | | | | | | X | X | X | X | |
| Record/reports AEs | | X | X | X | X | X | X | X | X | X | X | X | X | X | X | X | X | X | X | X | X | X | X | X | X |

| | | | | | | | | | | | | | | | | | | | | | | | | | |
|---|---|---|---|---|---|---|---|---|---|---|---|---|---|---|---|---|---|---|---|---|---|---|---|---|---|
| Record/reports SAEs | | X | X | X | X | X | X | X | X | X | X | X | X | X | X | X | X | X | X | X | X | X | X | X | X |
| Begin AZITH | | | | | | | ? | ? | ? | ? | ? | ? | ? | ? | ? | ? | ? | ? | ? | | | | | | |

### Table S2. Study intervention and sample collections

| Study Samples | OPt | OPt | Inpatient/Challenge Unit | | | | | | | | | | | | | | | | | Outpatient (OPt) | | | | | |
|---|---|---|---|---|---|---|---|---|---|---|---|---|---|---|---|---|---|---|---|---|---|---|---|---|---|
| | SCR | Base-line | Check-in | Challenge | | | | | | | | | | | | | | | | Phone Calls | | | | | |
| **Visit** | **1-SCR** | **1-B** | **2** | **3** | **4** | **5** | **6** | **7** | **8** | **9** | **10** | **11** | **12** | **13** | **14** | **15** | **16** | **17** | **18** | **19** | **20-23** | **24** | **25** | **26** | **27** | **28** |
| **Day:** | **-30** | **-25** | **-1** | **0** | **1** | **2** | **3** | **4** | **5** | **6** | **7** | **8** | **9** | **10** | **11** | **12** | **13** | **14** | **15** | **16** | **17-20** | **21** | **28** | **35** | **42** | **56** |
| **Study intervention and samples** | | | | | | | | | | | | | | | | | | | | | | | | | | |
| Challenge | | | | (X) | | | | | | | | | | | | | | | | | | | | | | |
| NPA and/or nasal swab (or equivalent) | X | | X | | X | X | X | X | X | X | X | X | X | X | X | X | X | X | X | X | | X | X | X | X | X |
| NW | X | | X | | X | X | X | X | X | X | X | X | X | X | X | X | X | X | X | X | | X | X | X | X | X |
| Blood/ serum (10mL) | | | X | +4-6hrs | X | | X | | X | | X | | | X | | | | X | | | | X | X | X | X | X |
| Blood Innate/ Adaptive IR (2mL) | | X† | X | +4-6hrs | X | | X | | X | | X | | | X | | | | X | | | | X | X | | | X |
| Blood/PBMC (30mL or 40mL*) | | X† | X | +4-6hrs | X | | X | | X | | X | | | X | | | | X | | | | X* | X* | X* | X* | X* |
| T-OMICS WB (2.5mL) | | | X | +4-6hrs | X | | X | | X | | X | | | | | | | X | | | | | X | | | |
| Urine 25–50 mL | | | X | X | X | X | X | X | X | X | X | X | X | X | X | X | X | X | X | X | | X | X | X | X | X |
| Saliva (1–2 mL) | | | X | X | X | X | X | X | X | X | X | X | X | X | X | X | X | X | X | X | | X | X | X | X | X |

### Key for Tables S1 and S2

OPt = Outpatient
MH = Medical history
Demo = Demographics
EKG = Electrocardiogram
PE = Physical exam
HBV and HCV = Hepatitis B virus and hepatitis C virus
DCs = Diary cards
AEs = Adverse events
SAEs = Serious adverse events
AZITH = Azithromycin
NPS = Nasopharyngeal swab
NPA = Nasopharyngeal aspirate
NW = Nasal wash
Blood/Serum is collected in non-gel serum separation tubes
IR = Immune Response
Blood Innate/Adaptive IR was collected in EDTA blood tubes
Blood/PBMC was collected in ACD blood tubes
T-OMICS WB = Transcriptomics from whole blood, collected in PAXgene tubes

- Inpatient period could be extended or shortened as per Methods. Up to Day 56 is shown, but participants were monitored up to one year from challenge Day.
- Challenge: Intranasal inoculation with a predetermined dose of *B. pertussis*.
- Study participation started from screening. Potentially eligible volunteers provided written informed consent prior to any screening procedure. The screening window was 30 days.
- Screening: Obtained written informed consent, reviewed eligibility criteria, reviewed and recorded medical history, reviewed and recorded demographics, EKG (baseline and screen for QT prolongation), SARS-CoV-2 screening by PCR from NP swab sample, pertussis serology screening.
- At Day -1, baseline SARS-CoV-2 serology was conducted.
- Medications: Recorded prior and concomitant medications and vaccinations to Day 28; on study Days 35, 42, 56 recorded only antibiotics and vaccines.
- Physical examination: Full physical examination on Check-In Day and Challenge Day, followed by delegated daily general exams during the inpatient period. In the event of an AE or SAE, a symptom-directed physical examination may have been conducted.
- Follow-up outpatient visits: History/symptom-directed examination.
- Follow-up phone calls (post-discharge): For symptomatic participants, asked about symptoms. For asymptomatic participants treated on Day 16, confirmed azithromycin daily dose is taken at the same time point each day as per instruction upon discharge, and asked about occurrence of symptoms.
- Safety bloods: Blood for hematology, CBC with differential; blood for chemistry testing, creatinine, total bilirubin, BUN, ALT, AST, and CRP.
- Diary Card: Receipt of the DC on Day 16 (or discharge day); DCs may have been received earlier or later depending on their outcome and discharge day.
- Azithromycin treatment started 24–48 hours from onset of mild cough or on Day 16 for those who remained asymptomatic (shown as "?"), or later than Day 16 depending on outcome.
- EKG: On Screening Day, and also conducted upon completion of the 5-day azithromycin course (shown as "?").
- [†]Blood samples taken during screening visits for baseline innate/adaptive immune response and PBMCs were collected only from participants who were deemed eligible for the study.

**Table S3. Summary of participant disposition-exposed population**

| Participant Disposition | Total n (%) |
|---|---|
| Enrolled Population | 168 |
| Has the subject met all the study eligibility criteria? | |
| Yes | 100 |
| No | 68 |
| Confirm if participant is eligible to receive a challenge dose | |
| Yes | 79 |
| No | 21 |
| Exposed Population | 79 (100%) |
| Withdrew from study early | 18 (22.8%) |
| Primary reason for early termination | |
| SAE | 0 (0.0%) |
| Non-serious AE | 1 (1.3%) |
| Protocol Violation | 0 (0.0%) |
| Consent withdrawal, not due to an AE | 6 (7.6%) |
| Moved from the study area | 2 (2.5%) |
| Lost to follow-up | 8 (10.1%) |
| Study Termination | 0 (0.0%) |
| Other | 1 (1.3%) |

Notes: The number of subjects in exposed population is used as the denominator in percentage calculation. Enrolled population includes participants with consent.. n= number of participants

**Table S4. Vaccine priming status of participants for each dose administered**

| Dose (CFU) | aP-Primed<br>n (%) | wP-primed<br>n (%) |
|---|---|---|
| $10^4$ | 2 (33.3%) | 4 (66.7%) |
| $10^5$ | 0 (0%) | 5 (100%) |
| $5 \times 10^5$ | 0 (0%) | 5 (100%) |
| $10^6$ | 1 (16.7%) | 5 (83.3%) |
| $5 \times 10^6$ | 2 (20%) | 8 (80%) |
| $10^7$ | 11 (50%) | 11 (50%) |
| $5 \times 10^7$ | 4 (33.3%) | 8 (66.7%) |
| $10^8$ | 5 (55.6%) | 4 (44.4%) |
| Total | 25 (33.3%) | 50 66.7%) |

**Table S5. Summary statistics for actual dose administered by dose group (exposed population: n=79)**

| Target Dose | Median Dose | Min Dose | Max Dose | Mean | Geometric Mean |
|---|---|---|---|---|---|
| $10^4$ | $1.0 \times 10^4$ | $0.84 \times 10^4$ | $1.1 \times 10^4$ | $0.96 \times 10^4$ | $0.96 \times 10^4$ |
| $10^5$ | $1.1 \times 10^5$ | $0.82 \times 10^5$ | $1.7 \times 10^5$ | $1.1 \times 10^5$ | $1.1 \times 10^5$ |
| $5 \times 10^5$ | $5.7 \times 10^5$ | $3.4 \times 10^5$ | $8.2 \times 10^5$ | $5.5 \times 10^5$ | $5.2 \times 10^5$ |
| $10^6$ | $1.0 \times 10^6$ | $0.58 \times 10^6$ | $1.1 \times 10^6$ | $0.95 \times 10^6$ | $0.93 \times 10^6$ |
| $5 \times 10^6$ | $7.1 \times 10^6$ | $4.4 \times 10^6$ | $\underline{13.0 \times 10^6}$ | $7.8 \times 10^6$ | $7.3 \times 10^6$ |
| $10^7$ | $1.0 \times 10^7$ | $0.68 \times 10^7$ | $1.9 \times 10^7$ | $1.1 \times 10^7$ | $1.0 \times 10^7$ |
| $5 \times 10^7$ | $6.4 \times 10^7$ | $2.0 \times 10^7$ | $\underline{11.0 \times 10^7}$ | $6.9 \times 10^7$ | $6.3 \times 10^7$ |
| $10^8$ | $1.1 \times 10^8$ | $0.84 \times 10^8$ | $2.3 \times 10^8$ | $1.3 \times 10^8$ | $1.2 \times 10^8$ |

Notes: Underlined indicates the doses where the maximum dose administered in a study group overlapped with the next higher target dose.

**Table S6. Symptomatic disease, and symptomatic disease with cough, for the stage-two analysis**

| Dose (CFU) | n Total Exposed | n, (%) of Total Exposed | | n, (%) of Total Symptomatic |
|---|---|---|---|---|
| | | Total symptomatic | Symptomatic with cough | Symptomatic with cough |
| $5\times10^6$ | 10 | 2 (20%) | 1 (10%) | 1 (50%) |
| $10^7$ (HID70-90) | 22 | 16 (72.7%) | 8 (36.4%) | 8 (50%) |
| $5\times10^7$ | 12 | 7 (58.3%) | 3 (25.0%) | 3 (42.9%) |
| $10^8$ | 9 | 5 (55.6%) | 3 (33.3%) | 3 (60%) |
| Total | 53 | 30 | 15 | 15 |

**Table S7. The percent seroconversion for four pertussis antibodies by clinical outcome for the pooled four uppermost doses (stage-two analysis)**

| Outcome | No seroconversion<br>n (%) | Seroconversion<br>n (%) |
|---|---|---|
| Anti-PT IgG | | |
| Non-infected | 9 (100%) | 0 (0%) |
| Asymptomatic infection | 12 (85.7%) | 2 (14.3%) |
| Symptomatic infection | 14 (46.7%) | 16 (53.3%) |
| Anti-FIM IgG | | |
| Non-infected | 9 (100%) | 0 (0%) |
| Asymptomatic infection | 11 (78.6%) | 3 (21.4%) |
| Symptomatic infection | 24 (80.0%) | 6 (20.0%) |
| Anti-FHA IgG | | |
| Non-infected | 9 (100%) | 0 (0%) |
| Asymptomatic infection | 9 (64.3%) | 5 (35.7%) |
| Symptomatic infection | 11 (36.7%) | 19 (63.3%) |
| Anti-PRN IgG | | |
| Non-infected | 9 (100%) | 0 (0%) |
| Asymptomatic infection | 12 (85.7%) | 2 (14.3%) |
| Symptomatic infection | 21 (70.0%) | 9 (30.0%) |
| All | | |
| Non-infected | 9 (100%) | 0 (0%) |
| Asymptomatic infection | 9 (64.3%) | 5 (35.7%) |
| Symptomatic infection | 9 (30.0%) | 21 (70.0%) |

Notes: the percent seroconversion was determined relative to the n values per disease outcome: n=9 Non-infected, n=14, Asymptomatic infection, n=30 Symptomatic infection

**Table S8. The percent seroconversion for four pertussis antibodies by clinical outcome for all doses**

| Outcome | No seroconversion<br>n (%) | Seroconversion<br>n (%) |
|---|---|---|
| Anti-PT IgG | | |
| Non-infected | 25 (100%) | 0 (0%) |
| Asymptomatic infection | 13 (86.7%) | 2 (13.3%) |
| Symptomatic infection | 17 (48.6%) | 18 (51.4%) |
| Anti-FIM IgG | | |
| Non-infected | 25 (100%) | 0 (0%) |
| Asymptomatic infection | 12 (80%) | 3 (20%) |
| Symptomatic infection | 29 (82.9%) | 6 (17.1%) |
| Anti-FHA IgG | | |
| Non-infected | 25 (100%) | 0 (0%) |
| Asymptomatic infection | 10 (66.7%) | 5 (33.3%) |
| Symptomatic infection | 14 (40.0%) | 21 (60%) |
| Anti-PRN IgG | | |
| Non-infected | 25 (100%) | 0 (0%) |
| Asymptomatic infection | 13 (86.7%) | 2 (13.3%) |
| Symptomatic infection | 25 (71.4%) | 10 (28.6%) |
| All | | |
| Non-infected | 25 (100%) | 0 (0%) |
| Asymptomatic infection | 10 (66.7%) | 5 (33.3%) |
| Symptomatic infection | 12 (34.3%) | 23 (65.7%) |

Notes: the percent seroconversion was determined relative to the n values per disease outcome: n=25 Non-infected, n=15 Asymptomatic infection, n=35 Symptomatic infection.

**Table S9. Summary of unsolicited adverse events - exposed population**

| Unsolicited AE Details | Total (N=79) n (%), E |
|---|---|
| At least one unsolicited AE | 59 (74.7%), 181 |
| At least one >= Grade 3 Unsolicited AE | 7 (8.9%), 10 |
| At least one Related Unsolicited AE | 3 (3.8%), 3 |
| At least one >= Grade 3 Related Unsolicited AE | 0 (0.0%), 0 |
| At least one Unsolicited SAE | 1 (1.3%), 3 |
| At least one Related Unsolicited SAE | 0 (0.0%), 0 |
| At least one >= Grade 3 Related Unsolicited SAE | 0 (0.0%), 0 |
| At least one unsolicited AE leading to inpatient hospitalization or prolongation of existing hospitalization | 1 (1.3%), 2 |
| At least one unsolicited AE leading to death | 0 (0.0%), 0 |
| At least one unsolicited AE leading to life threatening | 0 (0.0%), 0 |
| At least one unsolicited AE leading to persistent or significant disability/incapacity | 1 (1.3%), 2 |
| At least one unsolicited AE leading to a congenital anomaly/birth defect | 0 (0.0%), 0 |
| At least one unsolicited AE leading to other important medical event | 0 (0.0%), 0 |
| At least one unsolicited AE leading to concomitant medications | 29 (36.7%), 53 |
| At least one unsolicited AE leading to medical procedures | 1 (1.3%), 2 |
| At least one unsolicited AE leading to withdrawal from study | 1 (1.3%), 1 |

n (%), E: n = Count of Participants (at least one event i.e. Participants counted only once if the participant reported one or more Events), % = (n / Number of Participants in Exposed Population) *100, E = Count of Events (Participant may be counted more than once). Unsolicited AEs also include SAEs.

**Table S10. Proportion of negative culture results during azithromycin therapy for those infected at start of treatment**

| Days post-treatment | Proportion of negative culture results by NW and/or NPA |
|---|---|
| 0 | 0 (0.0%) |
| 1 | 17 (41.5%) |
| 2 | 31 (75.6%) |
| 3 | 38 (92.7%) |
| 4 | 40 (100%) |

Notes: NPA= Nasopharyngeal aspirates. NPAs were included in this assessment with nasal washes (NW) (Table S2). Excludes those that had cleared infection before the time of treatment.

**Figures S1. GLMM analysis**

**Below are proportion plots of symptom reporting alongside the generalized linear mixed model (GLMM) plot for each of the reported nine solicited symptoms. Both the non-infected and symptomatic infection groups are shown per figure.**

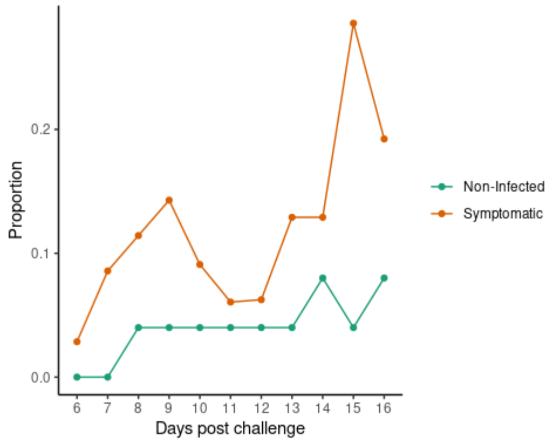

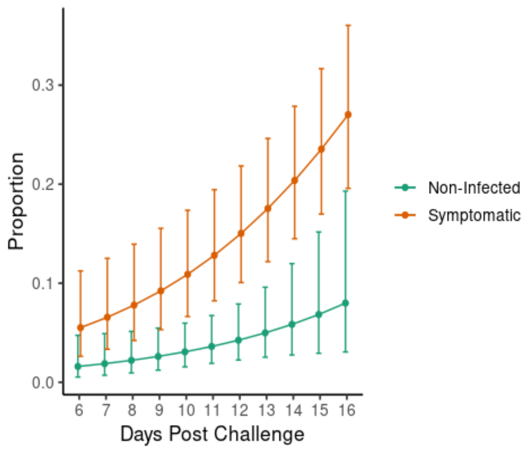

**A. Malaise**

The slope of days post challenge is 0.71 in the non-infected group compared to a slope of 1.18 in the symptomatic infection group.

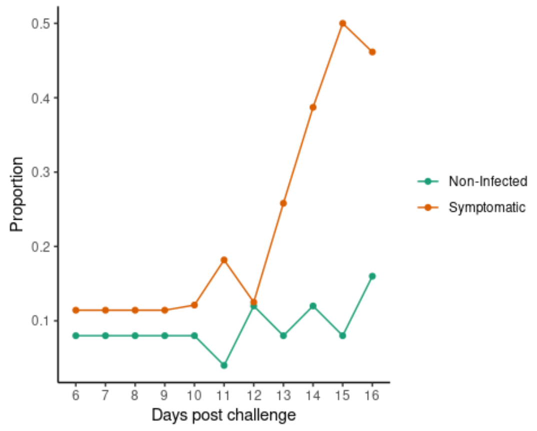

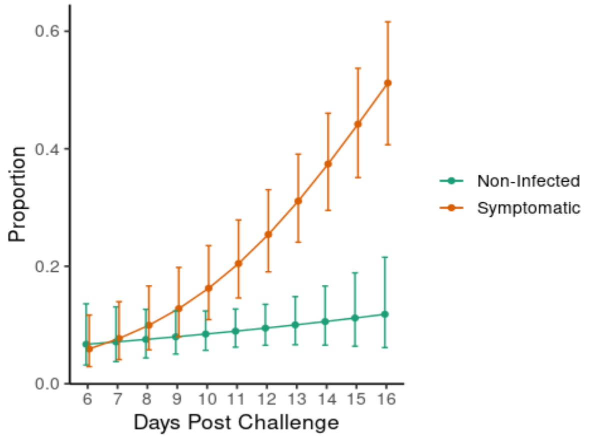

**B. Fatigue**

The slope of days post challenge is 0.23 in the non-infected group compared to a slope of 1.36 in the symptomatic infection group.

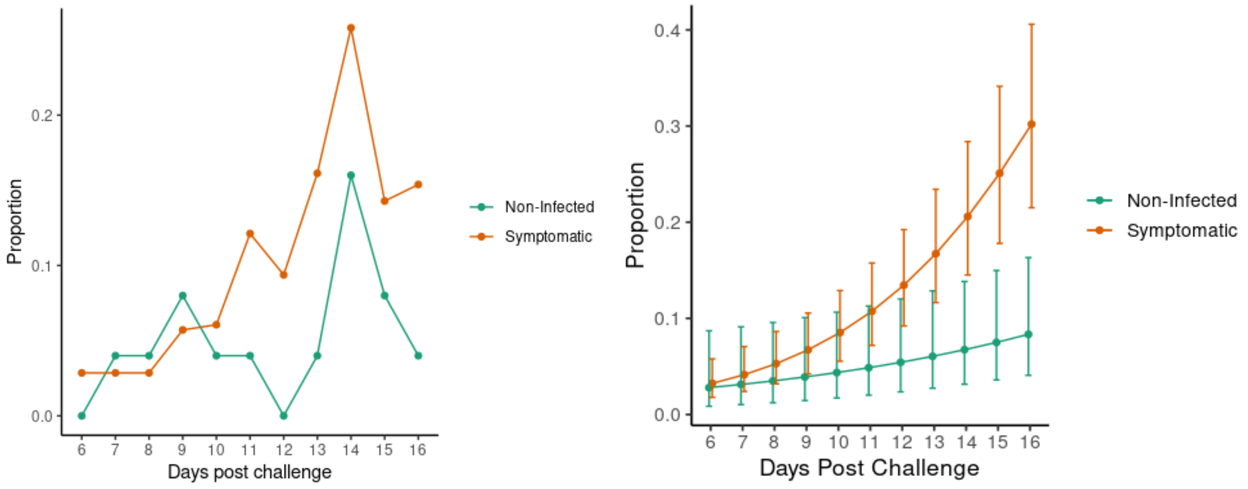

symptomatic and non-infected

**C. Runny nose**

The slope of days post challenge is 0.74 in the non-infected group compared to a slope of 1.44 in the symptomatic infection group.

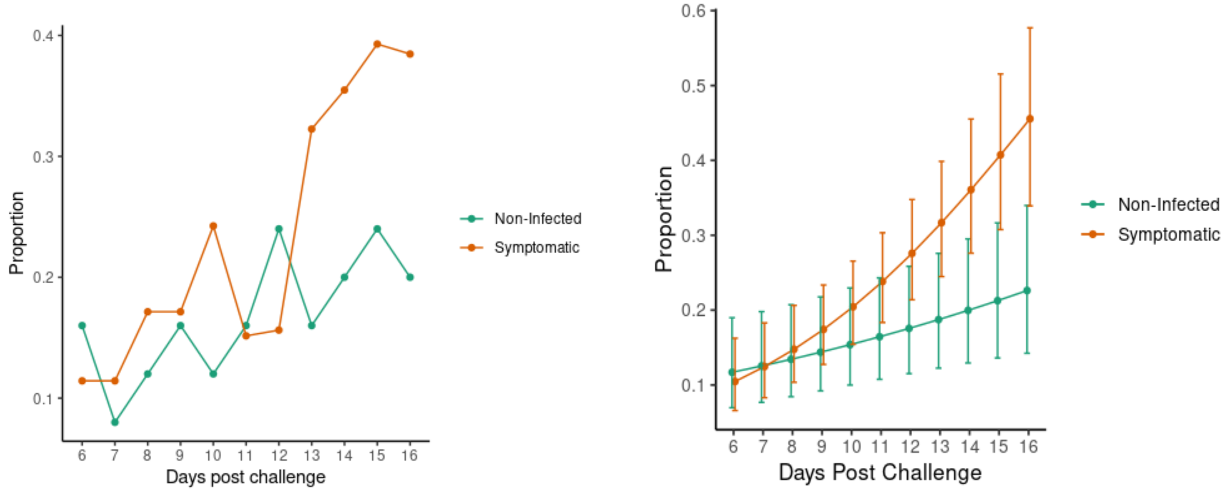

**D. Nasal Congestion**.

The slope of days post challenge is 0.43 in the non-infected group compared to a slope of 0.94 in the symptomatic infection group.

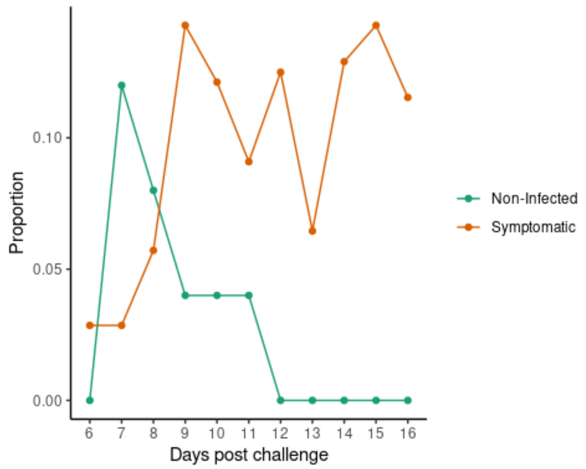

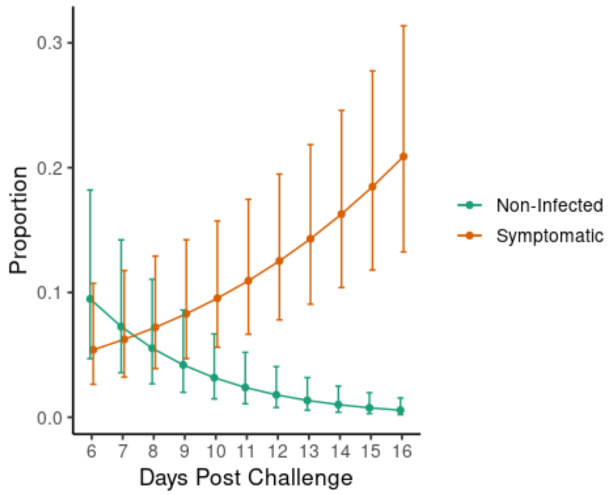

**E. Sneezing**

The slope of days post challenge is -1.53 in the non-infected group compared to a slope of 1.42 in the symptomatic infection group.

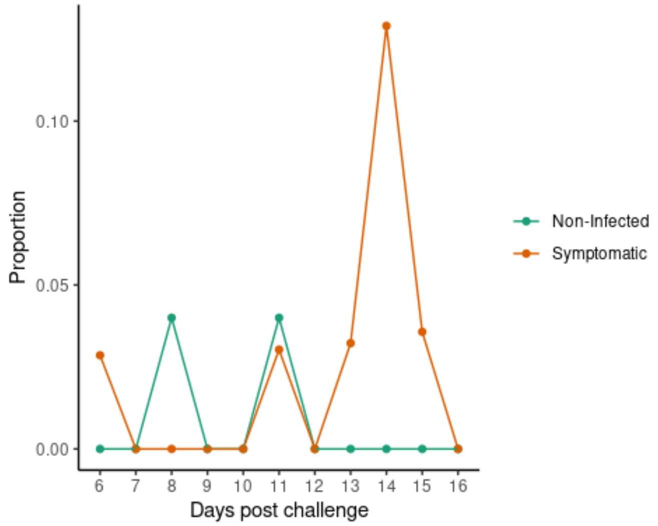

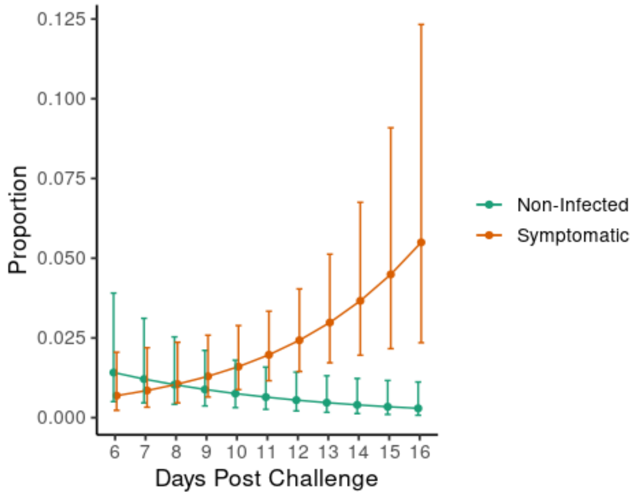

**F. Watery eyes**

The slope of days post challenge is -0.55 in the non-infected group compared to a slope of 1.13 in the symptomatic infection group.

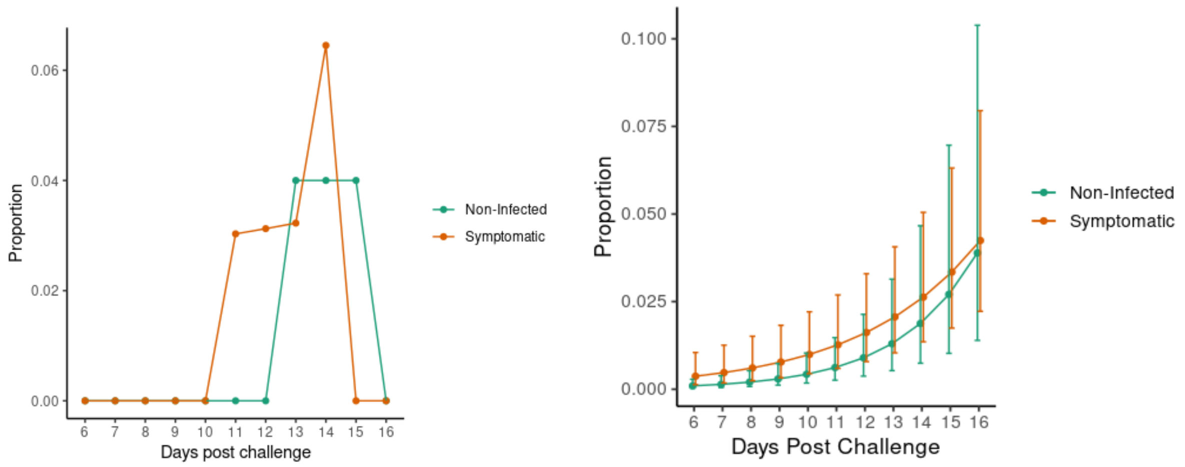

### G. Red eyes

The slope of days post challenge is 1.58 in the non-infected group compared to a slope of 2.15 in the symptomatic infection group.

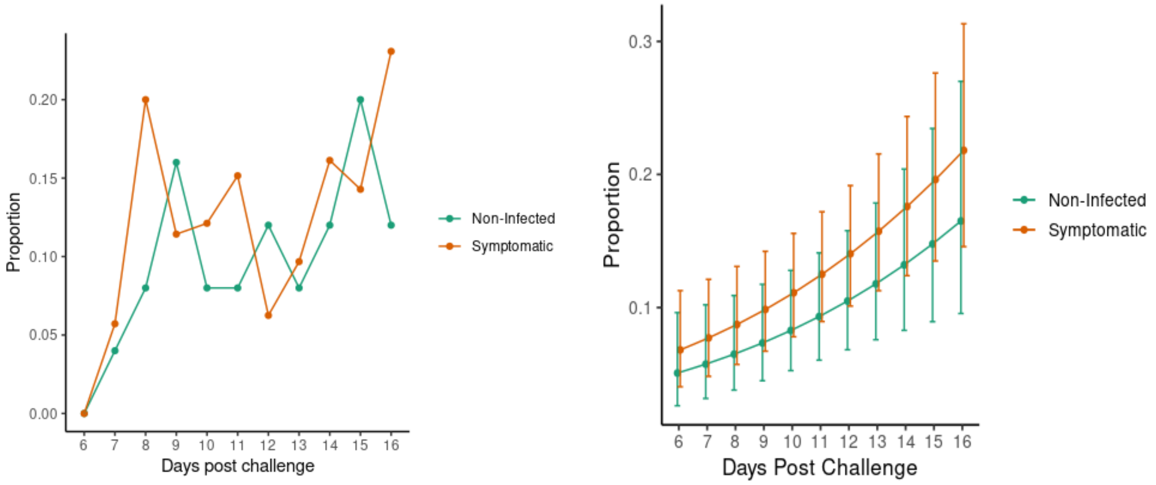

### H. Sore throat

There is an effect of days post challenge (the effect is the same in both groups). The slope is 0.55.

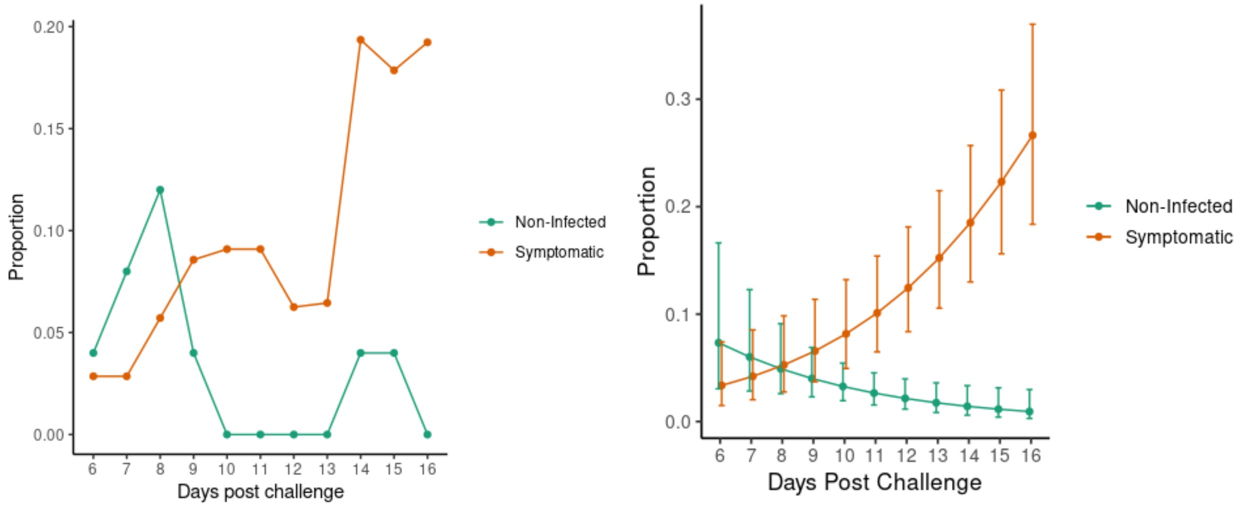

**I. Cough**

The slope of days post challenge is -0.81in the non-infected group compared to a slope of 2.34 in the infected group.

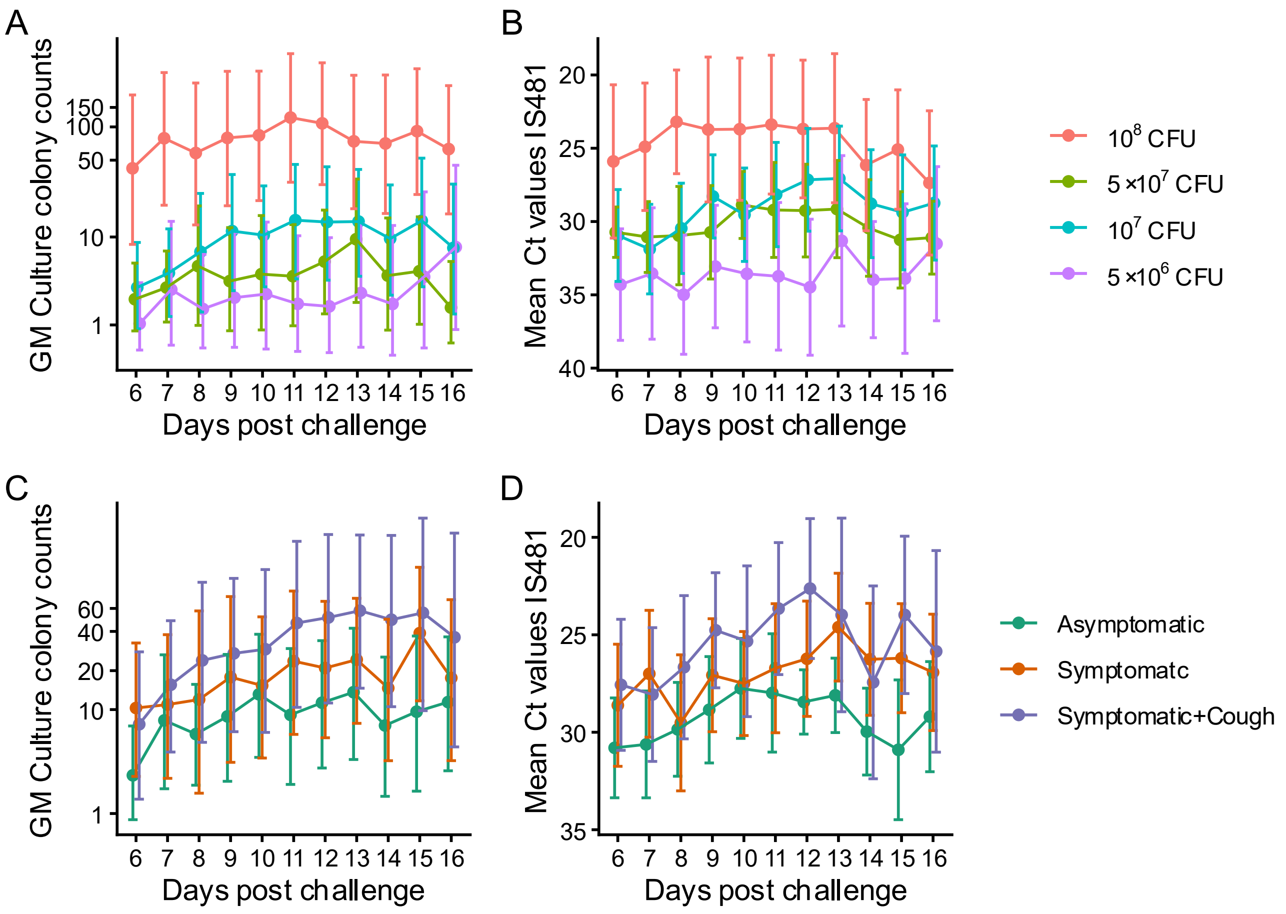

.

**Figures S2.** ***B. pertussis*** **detection in nasal wash over time post-challenge**

*B. pertussis* detection (average) in nasal wash over time post-challenge for the four (stage-two) study doses: A) geometric mean (GM) colony counts by days; B) mean Ct IS*481* values by days; C) GM colony counts by asymptomatic infection, symptomatic infection, and symptomatic infection with cough and; D) mean Ct IS*481*values by asymptomatic infection, symptomatic infection, and symptomatic infection with cough. Confidence intervals (95%) are shown.

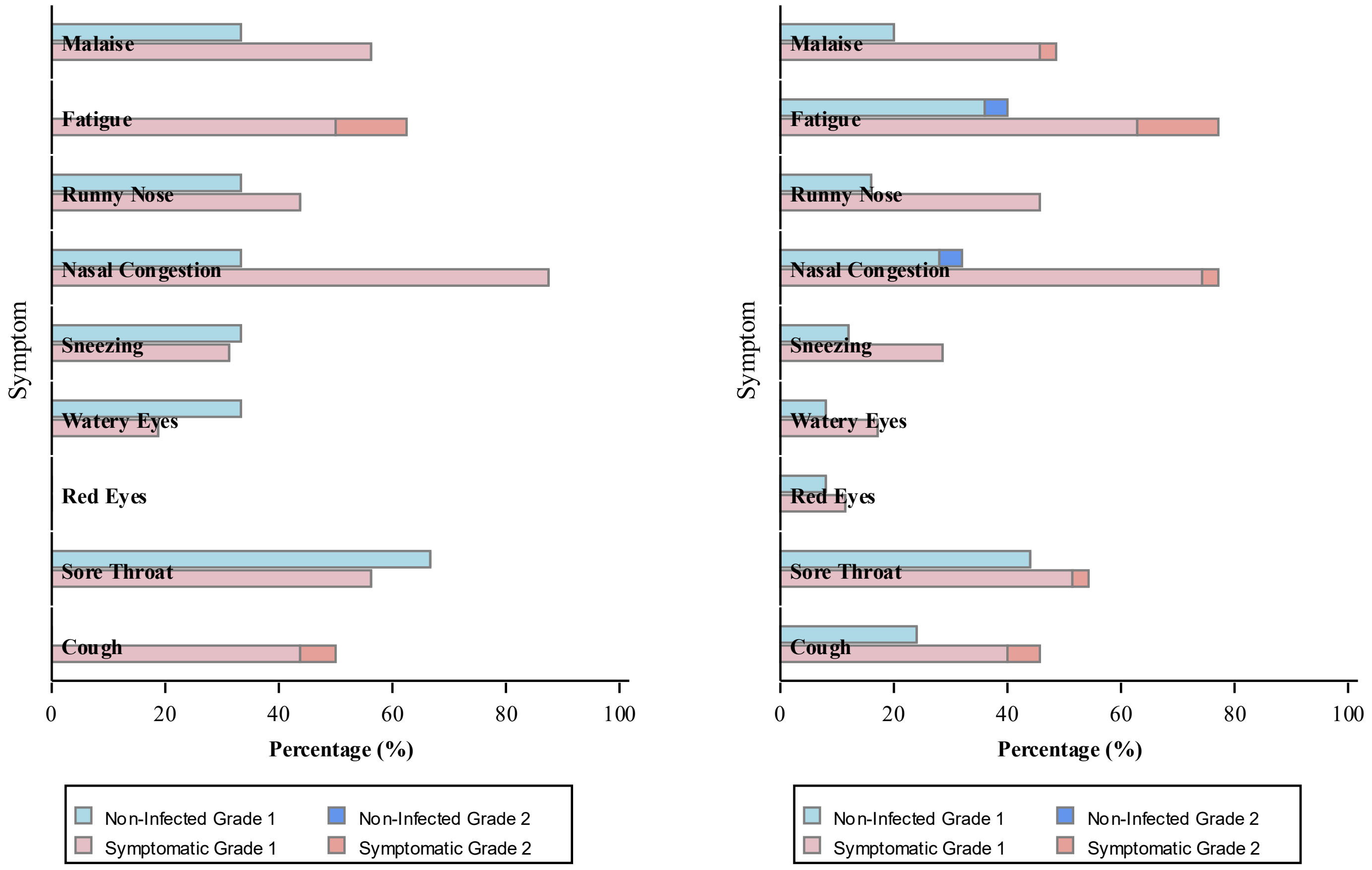

A. $10^7$ CFU

B. All doses

**Figures S3. Solicited symptoms for non-infected and symptomatic infection**

Percentage of participants reporting each solicited symptom for non-infected and symptomatic infection. A) HID70-90 (107 CFU) and B) all study doses.

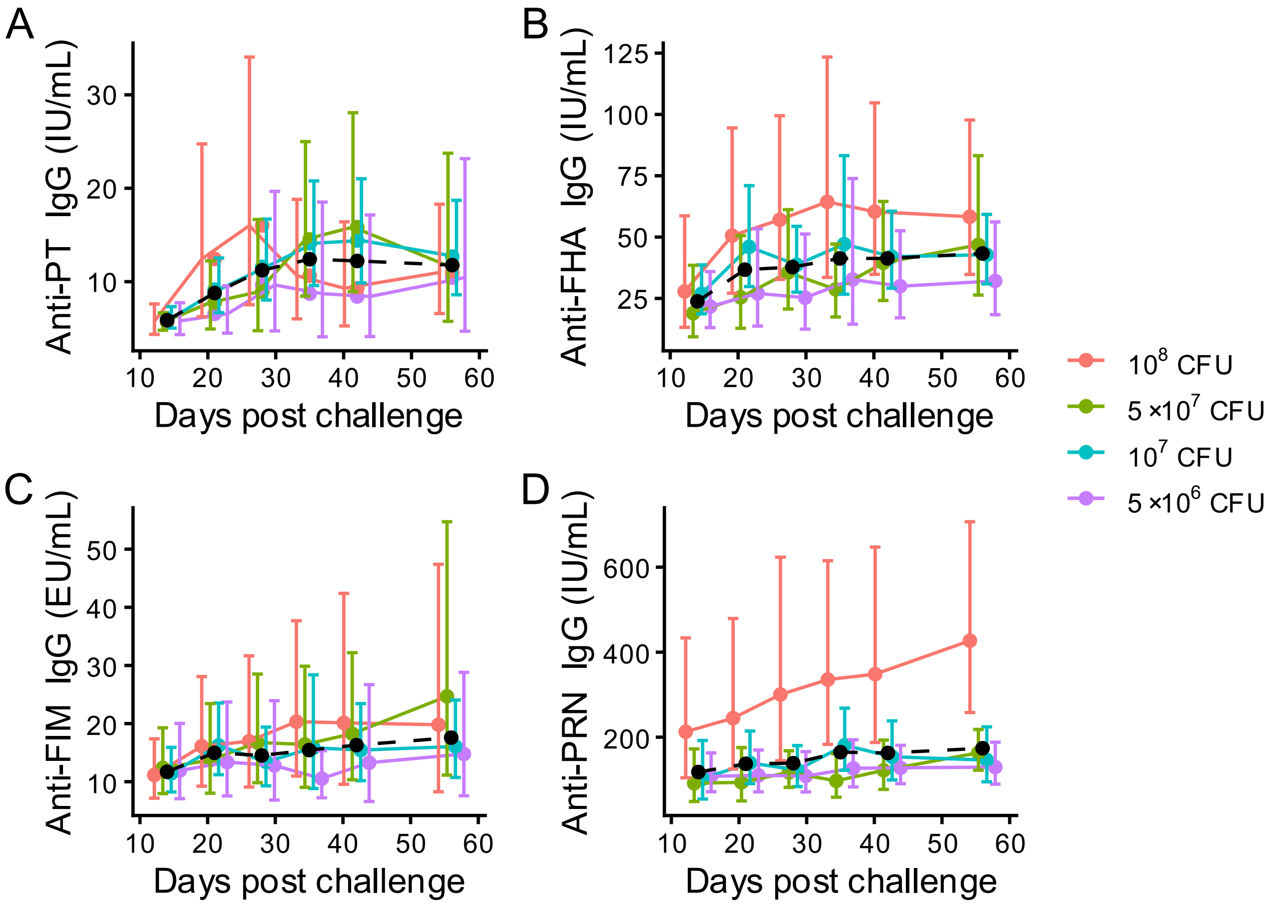

**Figures S4. Geometric mean titres over time post-challenge for pertussis antibodies**

Geometric mean titres over time post-challenge for pertussis antibodies at the four uppermost (stage-two) doses. A) anti-PT IgG; B) anti-FHA IgG; C) anti-FIM IgG; and D) anti-PRN IgG. The dotted line indicates the average among groups. Confidence intervals (95%) are shown.

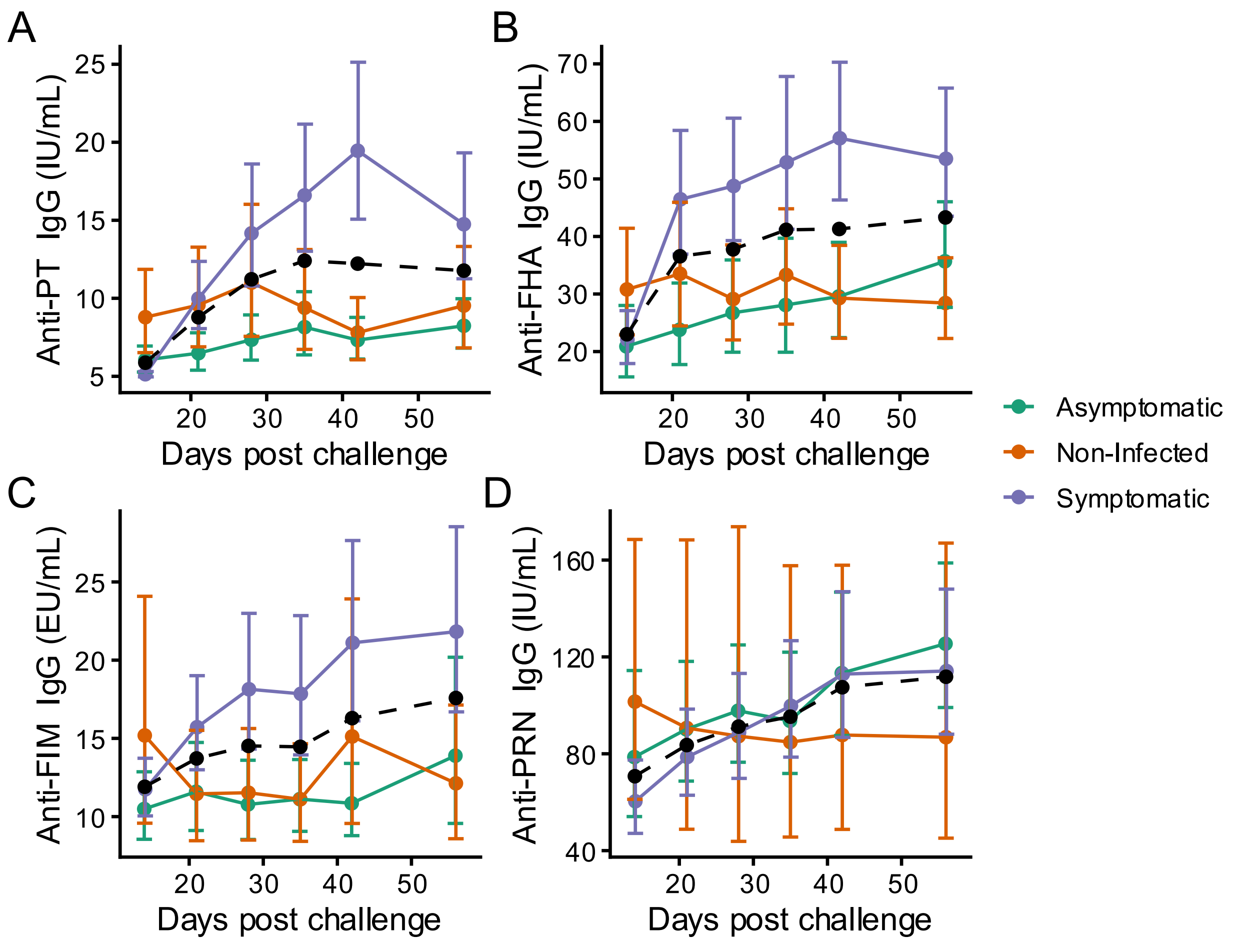

**Figures S5. Geometric mean titres for the pertussis antibodies by clinical outcome**

Geometric mean titres for the four pertussis antibodies pooled across four uppermost (stage-two) doses for each type of clinical outcome (symptomatic infection, asymptomatic infection and non-infected). A) anti-PT IgG; B) anti-FHA IgG; C) anti-FIM IgG; and D) anti-PRN IgG. The dotted line indicates the average among groups. Confidence intervals (95%) are shown.

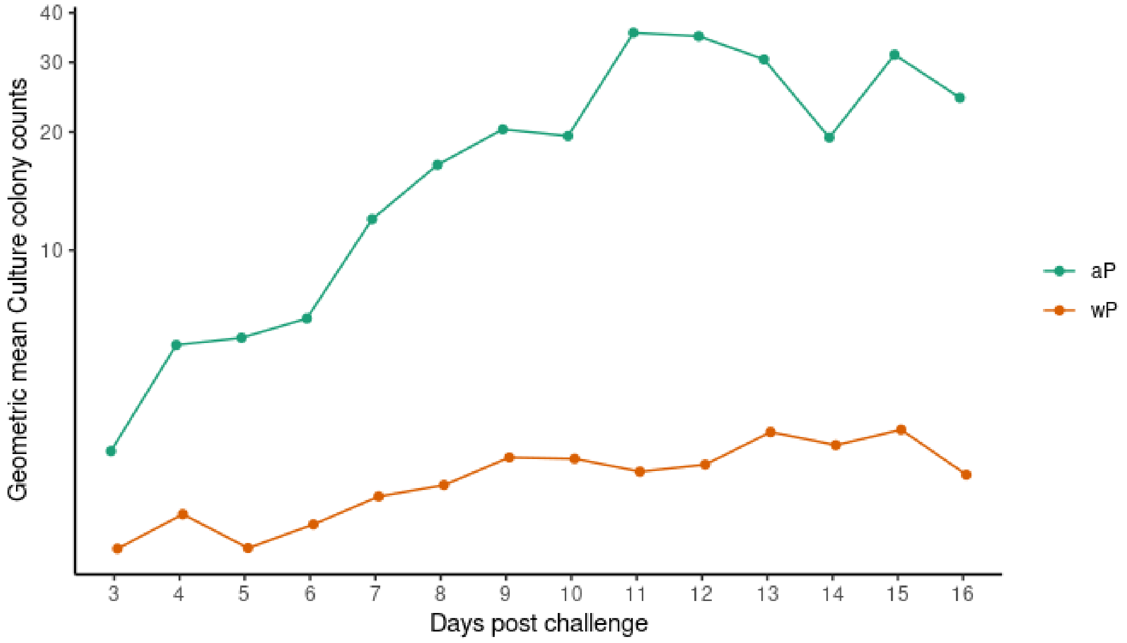

**A.** Geometric mean culture colony counts post-challenge by vaccine priming status (aP n=25; wP n=50) using nasal wash

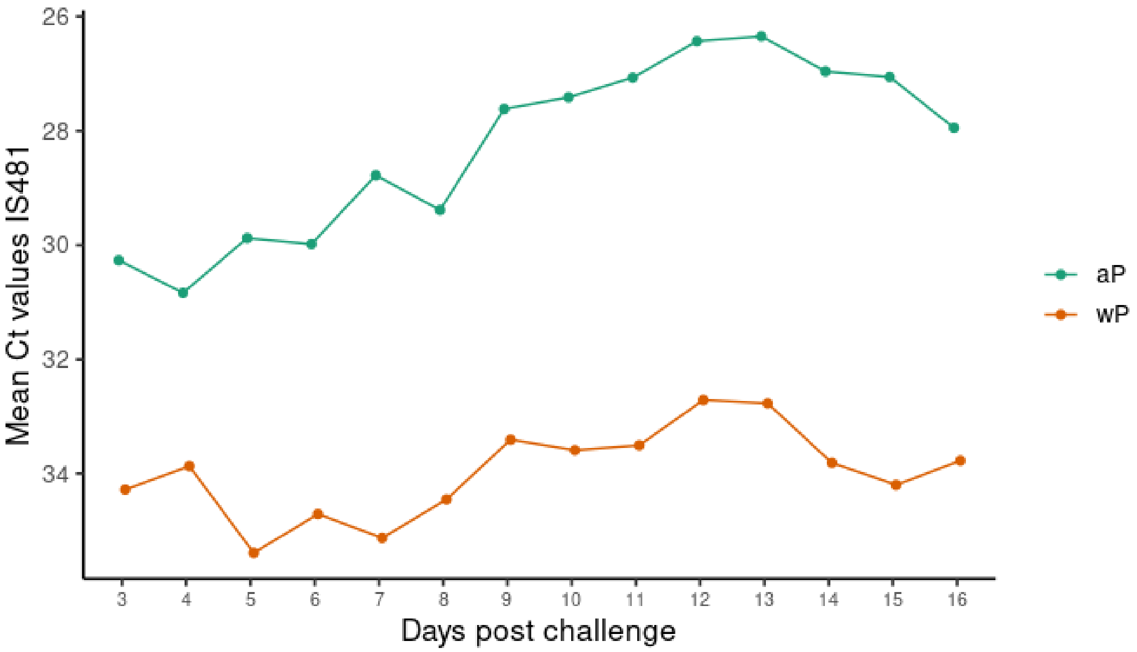

**B.** PCR Ct values post-challenge by vaccine priming status (aP n=25; wP n=50) using nasal wash.

**Figures S6. Vaccine priming and *B. pertussis* load**

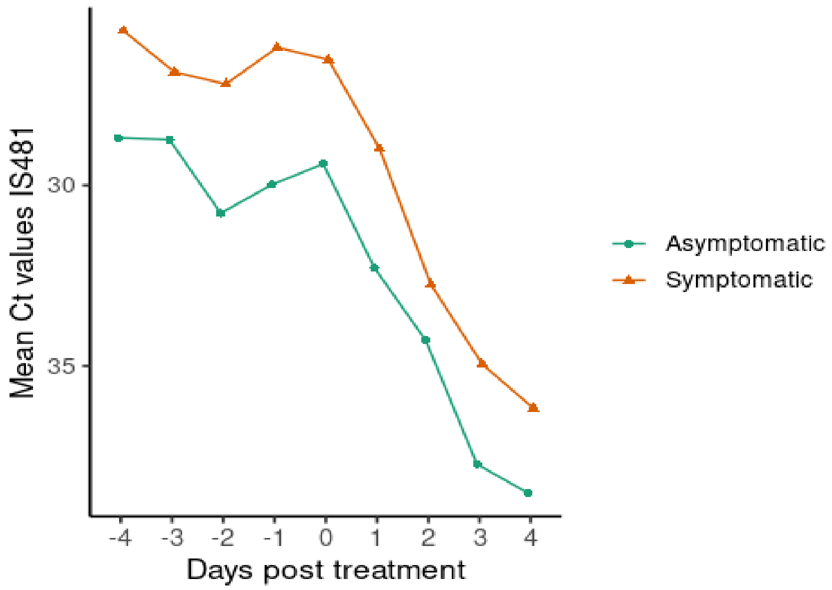

**A.** *B. pertussis* detection during azithromycin treatment over time (PCR of nasal wash) for symptomatic (n=35) and asymptomatic (n=15) participants

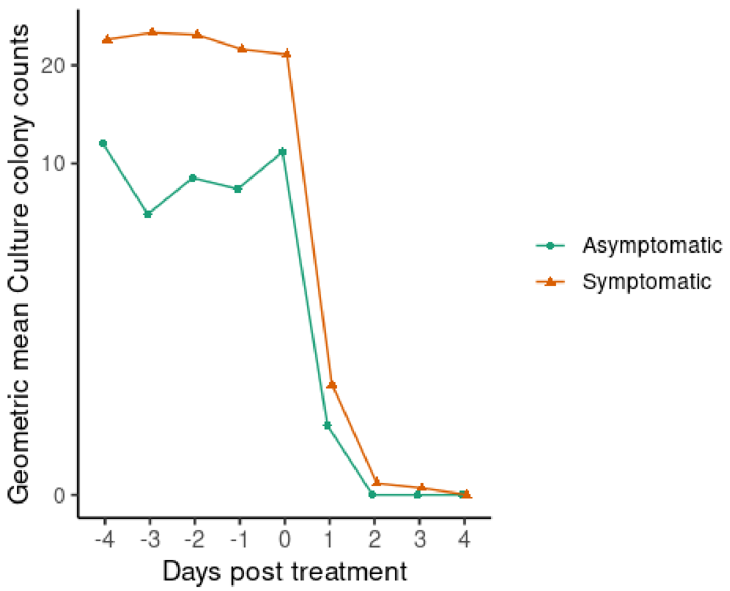

**B.** *B. pertussis* detection during azithromycin treatment over time (culture of nasal wash) for symptomatic (n=35) and asymptomatic (n=15) participants

**Figures S7. *B. pertussis* load pre- and post-treatment**

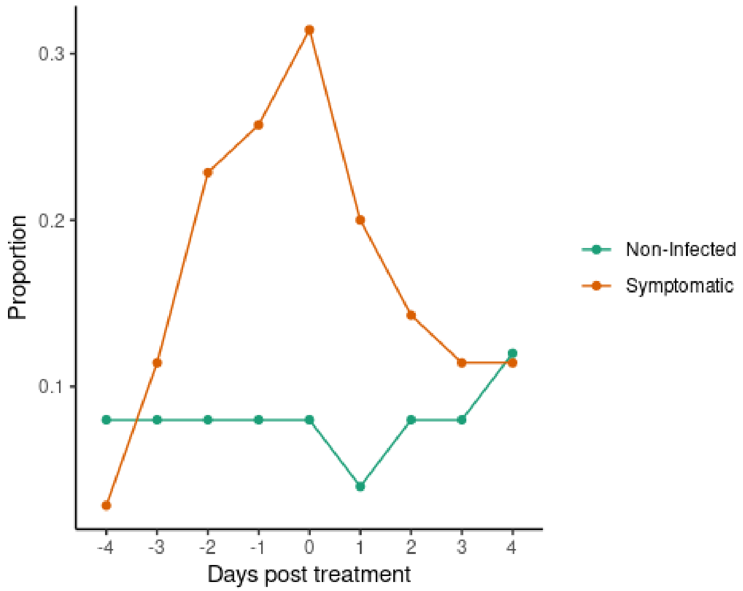

**Figures S8. Cough frequency pre- and post-treatment**

Proportion of symptomatic (n=35) and non-infected (n=25) participants with a cough before and after azithromycin treatment. Treatment began on Day 0 and continued up until Day 4.
